# The Protective Effect of Virus Capsids on RNA and DNA Virus Genomes in Wastewater

**DOI:** 10.1101/2023.05.19.23290245

**Authors:** Katherine R. Harrison, Delaney Snead, Anna Kilts, Michelle L. Ammerman, Krista R. Wigginton

## Abstract

Virus concentrations measured in municipal wastewater help inform both the water treatment necessary to protect human health and wastewater-based epidemiology. Wastewater measurements are typically PCR-based, and interpreting gene copy concentrations requires an understanding of the form and stability of the nucleic acids. Here, we study the persistence of model virus genomes in wastewater, the protective effects provided by the virus capsids, and the relative decay rates of genome and infectious viruses. In benchtop batch experiments at 25 °C, extraviral (+)ssRNA and dsDNA amplicons degraded by 90% within 15-19 minutes and 1.6-1.9 hours, respectively. When encapsidated, the T_90_ for MS2 (+)ssRNA increased by 424× and the T_90_ for T4 dsDNA increased by 52×. The (+)ssRNA decay rates were similar for a range of amplicon sizes. For our model phages MS2 and T4, the nucleic acid signal in untreated wastewater disappeared shortly after the viruses lost infectivity. Combined, these results suggest that most viral genome copies measured in wastewater are part of intact virus particles, that measured concentrations are independent of assay amplicon sizes, and that the virus genome decay rates of naked viruses are similar to inactivation rates. These findings will be valuable for the interpretation of wastewater virus measurements.

## INTRODUCTION

Many human viruses are present in municipal wastewater at high concentrations, and wastewater measurements have been widely used for tracking human diseases such as polio, COVID-19, influenza, respiratory syncytial virus (RSV), norovirus, Mpox, and other enteric and non-enteric viruses.^1–22^ Beyond wastewater-based epidemiology (WBE) applications, wastewater virus measurements are also critical for determining the extent of advanced water treatment that is necessary in water reuse applications to achieve acceptable levels of risk in the finished drinking water.^23–26^ Quantifying viruses in wastewater most frequently involves molecular-based methods (i.e. quantitative PCR methods) due to the limitations and complexities of culture-based methods. PCR-based quantification typically targets small portions of the viral genome and therefore the resulting concentrations are likely higher than infectious virus particles. The interpretation and application of gene copy concentrations requires an understanding of the genome signal stability as wastewater is conveyed through the sewage system, and of the relative concentrations of genome copies and infectious virus particles.

Most human viruses enter the wastewater through urine and stool. Some viruses, such as human norovirus and human adenoviruses, are excreted in stool as infectious virions at extremely high concentrations.^27^ Other viruses, such as enveloped SARS-CoV-2, are excreted primarily in a non-infectious state.^28,29^ Furthermore, due to the fact that many RNA viruses generate extensive amounts of subgenomic mRNA sequences as they replicate in the cytoplasm (i.e. extraviral nucleic acids and membrane-bound vesicles),^30–32^ these extraviral nucleic acids or membrane-bound vesicles are likely excreted and enter sewage systems. After entering the sewage system, wastewater spends an average of 3.3 hours and a maximum of 36 hours in a domestic sewer system before it enters a wastewater treatment plant,^33^ with travel times decreasing during wet-weather conditions.^34^ During that time, the integrity of intact virions can be impacted by changes in wastewater pH and temperature,^35,36^ chemical wastewater components such as surfactants,^37^ and biological components such as protists,^38^ and proteases.^39^ Based on these factors, the viral genomes detected in wastewater may be part of an infectious virus particle, an intact viral particle that is no longer infectious, a compromised virus particle, or as extraviral nucleic acids. The relative stability of the different states of viral nucleic acids in wastewater remains poorly characterized. Consequently, it is not clear what proportion of viral genes in wastewater samples are part of intact virus particles and how those genes have decayed as they are transported in wastewater.

Accurate genome decay rate constants in wastewater are important for linking gene copy concentrations measured at wastewater treatment plants to the number of gene copies excreted in the sewershed and overall community prevalence. For example, some prevalence-based models assume that SARS-CoV-2 RNA decay in the sewershed prior to sampling is of similar magnitude to PMMoV RNA decay.^3,40^ Other models have used first order decay rate constants in the range 0.1-1 days^-1^ for SARS-CoV-2 RNA to model community prevalence or infection rates.^34^ Beyond SARS-CoV-2, there is limited research on the relative stability of different viral nucleic acids in wastewater. In particular, there is a need for data on a broader range of nucleic acid types.

Different labs use a range of PCR-based methods for detecting the same viruses in wastewater, and the assays can target unique genome regions and amplicon sizes. Even within the same lab there can be a range of amplicon sizes employed to target different variants of the same virus sequence. For example, amplicons with a 2.5-fold difference in length (77-200 bases) were used to target different SARS-CoV-2 variants in one study.^41^ Previous research on genome amplicon sizes in water disinfection processes and environmental DNA (eDNA) persistence in fish species suggests that longer amplicons degrade faster than shorter amplicons.^42–44^ For example, when bacterial DNA was disinfected with free chlorine, amplicons that spanned 266-1017 bp exhibited up to a 2.4-fold difference in decay rate constants.^43^ A 2.6-fold difference was measured for the same amplicons during monochloramine disinfection.^43^ If the same amplicon size effect is true in wastewater, then the concentration of a pathogen measured in wastewater will be dependent on the amplicon size used in the PCR-based assay.

The purpose of this research is to characterize the persistence of viral (+)ssRNA and dsDNA in municipal wastewater, understand the protective effect of the virus capsid, and determine the potential biases of amplicon size on wastewater concentrations. We focus on (+)ssRNA and dsDNA viruses because the majority of human viruses found in municipal wastewater are Baltimore class I and IV viruses.^36^ We apply four model viruses, namely three bacteriophage and one coronavirus, with structures that are representative of typical human viruses in wastewater (Table 1). Ultimately, we compared virus infectivity and genome persistence in untreated wastewater to better define the relative persistence of genome signals and infectious viruses through experiments using one (+)ssRNA virus (MS2) and one dsDNA virus (T4). These detailed genome decay results will help in WBE applications, and the relative decay rates of infectious viruses and gene copies will help with the interpretation of gene copy concentration data in water reuse quantitative risk assessments.

**Table 1.** Characteristics of Viruses Used in This Study

| Virus | Genome type | Genome size | Capsid Structure | Particle size (nm) | Structure |
| --- | --- | --- | --- | --- | --- |
| MS2 | (+)ssRNA, linear | 3.6 kb | Icosahedral | ~25 | Nonenveloped |
| BCoV | (+)ssRNA, linear | 27-32 kb | Pleomorphic | 65-210 | Enveloped |
| T3 | dsDNA, linear | 38.2 kbp | Icosahedral head + tail | ~70 (head + tail) | Nonenveloped |
| T4 | dsDNA, linear | 168.9 kbp | Icosahedral head + tail | ~203 (head + tail) | Nonenveloped |

kb = kilobases, kbp = kilobase pairs

## MATERIALS AND METHODS

### Virus stocks, propagation, and plaque assays

Bacteriophage MS2, T3 and T4 were propagated and quantified with protocols that were published previously.^45,46^ Bacteriophage MS2 was propagated and titered with *E.coli* host ATCC #15597 and bacteriophages T3 and T4 were propagated and titered with *E. coli* host ATCC #11303. A summary of the media used for bacteriophage propagation and storage is provided in Supporting Information Table S1. Modified-live Bovilis Coronavirus Calf Vaccine (BCoV) was purchased from PBS Animal Health (Cat. No. 16445) and concentrated in TE Buffer (pH 8, Fisher Scientific) via centrifugal ultrafiltration with 10 kDa Amicon® centrifugal filter units. The MS2 stock was filter-sterilized through 0.22 μm polyethersulfone (PES) membrane filters (MilliporeSigma), and T3 and T4 stocks were filter-sterilized through 0.45 μm filters. The stock concentrations were ∼10^11^ gc mL^-1^, ∼10^9^ gc mL^-1^, and ∼10^10^ gc mL^-1^ for MS2, BCoV, and T3/T4, respectively. Bacteriophage MS2, T3, and T4 stocks were purified through a PEG-chloroform method described previously.^47^ Infectious MS2 and T4 bacteriophage were enumerated by plaque assay using the double agar overlay method.^45,46^ The virus stocks were stored in 1X PBS (Fisher) and kept at 4 °C prior to extraction. Bacteriophage stocks were titered at the start of each experiment.

### RNA and DNA extraction

Extraviral (+)ssRNA and dsDNA stocks were generated by extracting RNA and DNA from viral stocks immediately prior to experiments. (+)ssRNA was extracted from MS2 and BCoV virus stocks and dsDNA was extracted from T3 and T4 stocks using the AllPrep® PowerViral® DNA/RNA Kit (Qiagen) following the manufacturer’s protocol except that the bead beating step was not included. RNA was isolated from samples with beta-mercaptoethanol (Fisher). Samples were eluted into RNase-free water. The purity of the nucleic acid stocks was assessed with a NanoDrop Spectrophotometer (Thermo Scientific).

### Wastewater sampling and characterization

Wastewater influent was sampled from the City of Ann Arbor Wastewater Treatment Plant at the headworks post grit removal. The Ann Arbor Wastewater Treatment Plant has an average flow of 18.5 MGD and serves a population of 121,000. The 24-hour time-weighted composite samples (∼500 mL) were collected in autoclaved 1L bottles over an eleven-month period (November 2021 to October 2022) to capture seasonal variation in wastewater composition. All samples were transported to the University of Michigan campus and used in experiments immediately.

The wastewater sample pH was measured at the beginning and end of each experiment. The mean pH at the beginning of experiments was 7.74 and the mean decrease in pH during the course of the 7-12 day experiments was 0.95 (Table S2). The observed reductions in pH during experiments were likely due to biological activity that results in the formation of volatile fatty acids (VFAs) and thus lowers the pH of the wastewater. The wastewater sample temperature was maintained at 25 °C in a temperature-controlled room and the samples were gently mixed by continuous tilting on a rocking shaker (Fisher) to 30 degrees. It was previously estimated that 75% of global wastewater temperatures are within the range of 6.9–34.4 °C, so our experiments were conducted within this range.^64^ Total suspended solids (TSS) and total volatile suspended solids (TVSS) measurements were obtained from the Ann Arbor Wastewater Treatment Plant (Table S3). During this study, the mean TSS was 196 (± 51.4) mg L^-1^ and the mean TVSS was 166 (± 36.5) mg L^-1^.

### Wastewater experiments

Wastewater samples were spiked with either the intact viruses or the extracted viral nucleic acids; for the remainder of the manuscript, we defined these nucleic acids as either “encapsidated” or “extraviral”. The encapsidated virus stocks were treated with either RNase or DNase to confirm that the vast majority of the viruses were encapsidated (Figures S1 and S2). Likewise, extraviral dsDNA stocks were treated with DNase to confirm that the vast majority of these nucleic acids were extraviral and thus non-quantifiable after DNAse treatment (Figure S6). Details of these treatments are described in the Supporting Information. Control experiments were conducted in tandem in PBS solutions (10X PBS, pH 7.4 purchased from Fisher Scientific) to observe any background decay at room temperature.

For the extraviral nucleic acid experiments, 5 μL of freshly extracted MS2 (+)ssRNA, T3 dsDNA, and T4 dsDNA or 50 μL of freshly extracted BCoV (+)ssRNA was spiked into 500 μL of untreated wastewater. The MS2, T3 and T4 viruses were spiked into samples with a dilution factor of 1:100 to minimize any matrix effects from the extracted nucleic acid solution. BCoV was spiked into samples with a dilution factor of 1:10 due to the lower stock concentration. Extraviral nucleic acid experiments were performed in triplicate and each replicate was conducted in a different composite untreated wastewater sample collected on a different day. Extraviral nucleic acid was spiked into samples to achieve initial concentrations of ∼10^9^ gc mL^-1^ for MS2, ∼10^8^ gc mL^-1^ for BCoV, ∼10^8^ gc mL^-1^ for T3, and ∼10^8^ gc mL^-1^ for T4. The stock concentrations were determined by digital droplet polymerase chain reaction (ddPCR) assays. The quantified extraviral nucleic acids (Figure 1) were reported with respect to the expected initial concentrations to demonstrate the rapid decrease in (+)ssRNA concentrations in wastewater. Plots of encapsidated nucleic acids (Figure 2) were plotted with respect to the initial concentrations measured in samples immediately after spiking due to the fact that we could not confidently report the absolute abundance of encapsidated nucleic acids in the stocks. Issues with viral nucleic acid recoveries from stocks and controls have been documented in several other studies.^48,49^

**Figure 1.**
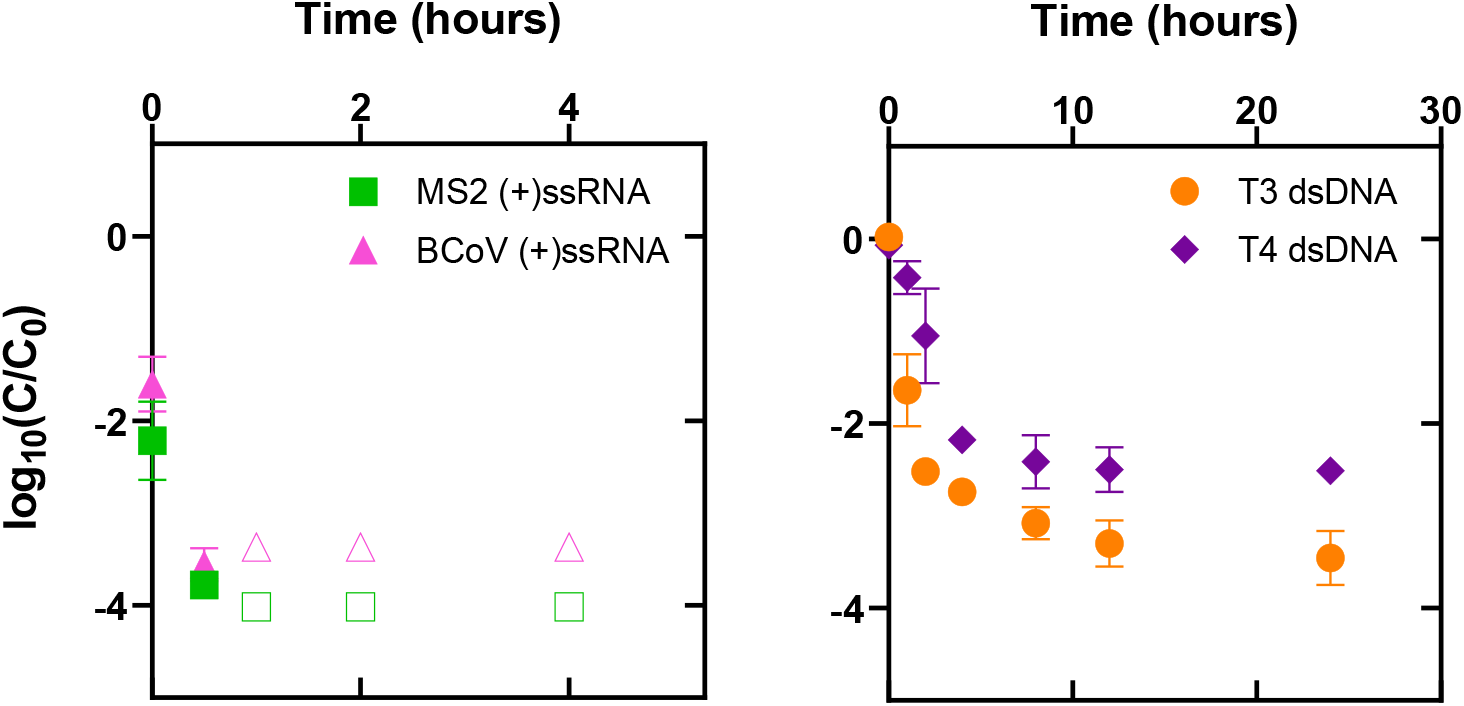
Extraviral (+)ssRNA and dsDNA decay in untreated wastewater at 25 °C. C represents the concentration of the viral nucleic acids in gene copies μL^-1^ at time t in hours and C_0_ represents the initial concentration of the viral nucleic acids at time = 0 based on the amount of stock that was spiked into the wastewater. Initial concentrations in wastewater were ∼10^9^ gc mL^-1^ for MS2 (+)ssRNA and ∼10^8^ gc mL^-1^ for BCoV (+)ssRNA, T3 dsDNA, and T4 dsDNA. Open symbols represent measurements that were below the limit of quantification and are plotted as the limit of quantification for each virus assay. Error bars represent the standard error for three experimental replicates conducted in wastewater samples collected on different days.

**Figure 2.**
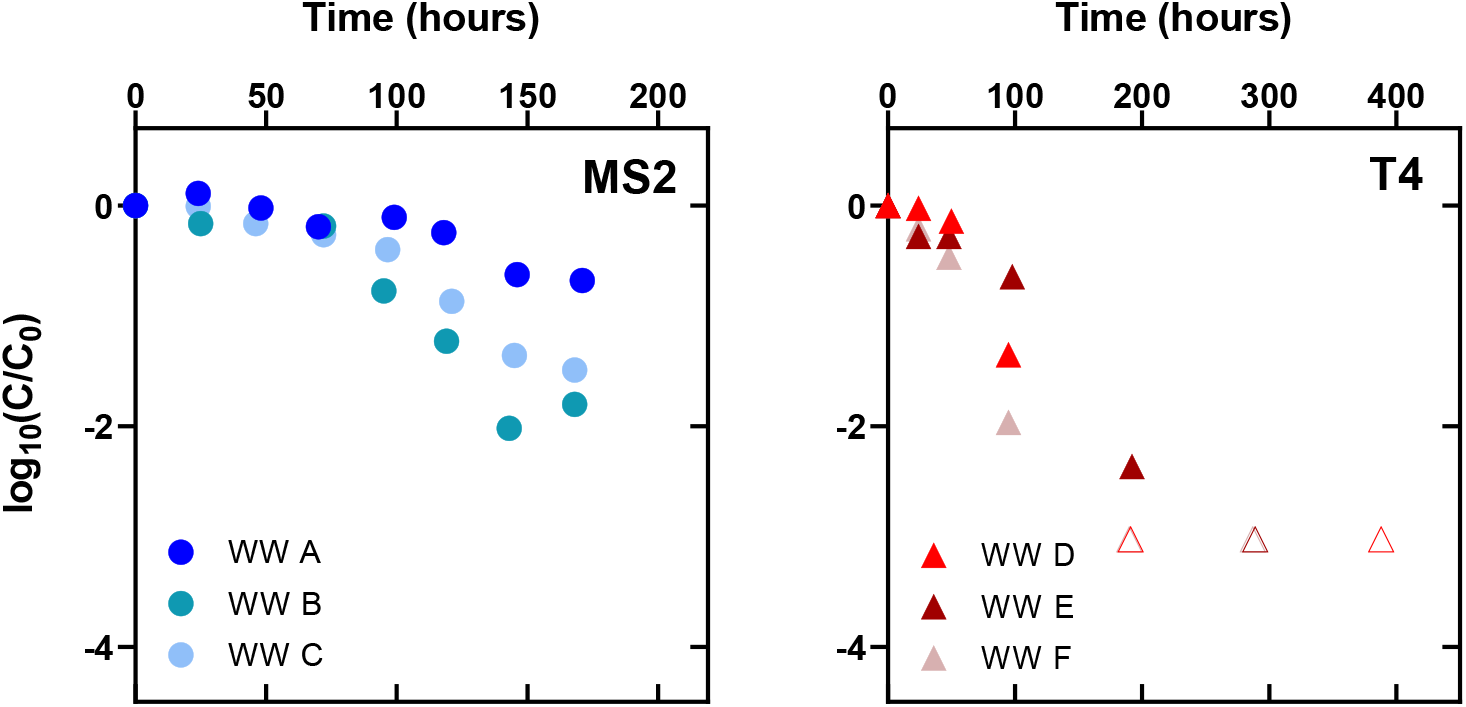
MS2 (+)ssRNA (left) and T4 dsDNA (right) encapsidated genome decay in untreated wastewater at 25 °C. C refers to the concentration of the viral nucleic acids in gene copies μL^-1^ at time t in hours. C_0_ represents the initial concentration of the viral nucleic acids at time = 0 based on the amount of gene copies measured immediately after the stocks were added to the sample. Initial concentrations in wastewater were ∼10^9^ gc mL^-1^ for MS2 and ∼10^8^ gc mL^-1^ for T4. Open symbols represent measurements that were below the limit of quantification and are plotted as the limit of quantification for each virus assay. Individual experimental replicates are shown for MS2 (wastewaters A, B, C) and T4 (wastewaters D, E, F).

For the encapsidated nucleic acid experiments and infectious virus experiments, a 30 μL aliquot of the MS2 or T4 stock was spiked into 30 mL of untreated wastewater, resulting in final virus concentrations of ∼10^8^ PFU mL^-1^ for MS2 and ∼10^7^ PFU mL^-1^ for T4. The starting concentrations were confirmed with plaque assays conducted immediately after the viruses were spiked into the samples. Control experiments were conducted in PBS. These experiments were performed in triplicate with each replicate experiment taking place on different days and in different wastewater samples. Aliquots were removed from the wastewater batch reactors at desired experimental timepoints for both nucleic acid quantification and infectious virus quantification. Nucleic acids were extracted from the aliquots as described above. For infectious virus quantification, samples were serially diluted in 10-fold increments prior to plating and plates were enumerated if they contained 2-250 plaques. Host control blanks and PBS blanks were plated regularly to rule out contamination of the host culture and agar as well as the PBS dilution buffer.

### Complementary DNA (cDNA) synthesis and molecular quantification

The (+)ssRNA extracts were converted to cDNA using iScript™ Advanced cDNA Synthesis Kit for RT-qPCR (Bio-Rad) following the manufacturer’s protocol. Each 20 μL reaction consisted of 4 μL 5x iScript Advanced Reaction Mix, 1 μL iScript Advanced Reverse Transcriptase, 2 μL RNA template, and 13 μL nuclease-free water. Reactions were briefly mixed and centrifuged and then run on a Mastercycler EP Gradient Thermal Cycler (Eppendorf) with a reverse transcription step for 20 minutes at 46 °C followed by a reverse transcriptase inactivation step for 1 minute at 95 °C. Following cDNA synthesis, samples were stored at -20 °C until analysis by qPCR or ddPCR. Assay information for qPCR and ddPCR as per MIQE guidelines^50^ is provided in Supporting Information Tables S4 and S5.

Gene targets for MS2 were designed in this study and varied in size from 99 bp to 395 bases. Targets for BCoV, T3, and T4 gene copy quantification were obtained from the literature and were on average ∼100 b/bp in size.^51–53^ We used DNA standards for qPCR and cDNA for RT-qPCR. The standards were prepared from freshly extracted virus stock during each experiment and were enumerated with ddPCR. Standard curves ranged from 10^2^−10^7^ gc μL^-1^ for MS2, 10^1^−10^5^ gc μL^-1^ for BCoV, and 10^1^−10^6^ gc μL^-1^ for T3 and T4. Triplicate non-template controls showed either no amplification or amplification below the limit of detection for the assay. Background controls consisted of extracted wastewater samples without spiked virus to assess the target abundance in native wastewater. Inhibition controls consisted of 1:10 dilution of the extracts. Background controls were below the limit of detection for all assays (Figure S5) and inhibition controls showed limited inhibition in the assays (Figure S4).

### MS2 Primer Design

To test the impact of amplicon length on the observed (+)ssRNA viral genome decay kinetics in untreated wastewater, we designed three nested primer sets with increasing amplicon lengths (99, 192, 395 bases) over the same region of the MS2 genome (see Supporting Information Table S4 for complete primer sequences and annealing temperatures). MS2 primer sets were designed using the complete MS2 genome from the NCBI GenBank Database (Accession number NC_001417). The primer sets were designed in Primer3 (version 4.1.0) with the following constraints: primer size range 18-23 bases, optimal primer melting temperature 60 °C, optimal primer GC content 50%, and zero self-complementarity. The product size ranges varied for each consecutive nested primer set using the same forward primer sequence for all primers. The designed primers were synthesized by Integrated DNA Technologies (IDT).

### Decay constants

First-order rate constants for viral nucleic acids and virus inactivation were determined according to the following equation:

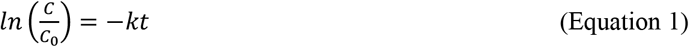

where t is the time when the sample is measured, C is the concentration of the targeted genome sequence (gene copies μL^-1^) or infectious virus (PFU mL^-1^) at time t, C_0_ is the concentration of the targeted genome sequence (gene copies μL^-1^) or infectious virus (PFU mL^-1^) at time 0, and k is the decay constant (time^-1^).

We calculated the mean rate constant and error after combining the three biological replicates in untreated wastewater into one dataset. When biphasic kinetics were observed, we calculated rate constants for the first phase and second kinetics phase.

T_90_, or the time it takes for 90% of the encapsidated virus genome to decay in untreated wastewater, was determined according to the following equation:

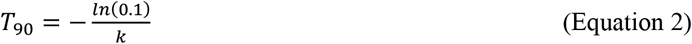

When biphasic kinetics were observed, T_90_ values were computed as the time when the virus concentration had decreased by 90% across both phases. An example calculation for T_90_ with biphasic kinetics is provided in the Supporting Information.

### Statistical analysis

All statistical analyses were performed with GraphPad Prism (Version 9.3.1).

## RESULTS AND DISCUSSION

### Extraviral RNA and DNA degradation kinetics

We compared the decay kinetics of extraviral nucleic acids from our four model viruses, MS2 (+)ssRNA, BCoV (+)ssRNA, T3 dsDNA, and T4 dsDNA (Table 2) in untreated wastewater at 25 °C. Each target amplicon was approximately 100 bases or base pairs. Both the (+)ssRNA and dsDNA nucleic acids underwent little decay in the PBS control samples (Figure S7). In wastewater, the extraviral (+)ssRNA degraded more rapidly than the extraviral dsDNA (Figure 1). Immediately after spiking the (+)ssRNA stocks into the wastewater, the measured extraviral MS2 and BCoV (+)ssRNA genome targets were already ∼2-log_10_ lower than the anticipated concentrations based on the volume of stock added. By contrast, the T3 and T4 dsDNA target concentrations were similar to the expected concentrations (∼10^8^ gc mL^-1^) immediately after the stocks were spiked into the wastewater samples. The MS2 and BCoV (+)ssRNA targets were no longer quantifiable after incubating in wastewater at room temperature for 30 minutes, corresponding to >3.8-log_10_ and >3.6-log_10_ decay for MS2 and BCoV (+)ssRNA, respectively. The T3 and T4 dsDNA targets were still quantifiable after 24 hours of incubation in wastewater but had decreased by 3.5-log_10_ and 2.5-log_10_ for T3 and T4 dsDNA, respectively.

**Table 2.** Summary of Kinetics Parameters (Mean ± 95% Confidence Interval) for Virus Genome Decay and Infectious Virus Decay in Untreated Wastewater at 25 °C

| | $k_{extraviral}$ (days<sup>-1</sup>) | | $T_{90}$ (time) | $k_{encapsidated}$ (days<sup>-1</sup>) | | $T_{90}$ (time) | $k_{infectious}$ (days<sup>-1</sup>) | | $T_{90}$ (time) |
| --- | --- | --- | --- | --- | --- | --- | --- | --- | --- |
|  | First phase: 0-0.5 hours |  |  | First phase:<br>0-72 hours | Second phase:<br>72-180 hours |  | First phase:<br>0-100 hours | Second phase:<br>100-180 hours |  |
| **MS2, 99 b** | $1.7 (\pm 0.76) \times 10^2$ | | 19 minutes | $1.7 (\pm 1.1) \times 10^{-1}$ | $6.9 (\pm 4.1) \times 10^{-1}$ | 5.6 days | $5.2 (\pm 1.9) \times 10^{-1}$ | $1.0 (\pm 0.90)$ | 4.3 days |
| **MS2, 192 b** | NA | | NA | $1.8 (\pm 1.9) \times 10^{-1}$<br>(NS) | $6.7 (\pm 7.0) \times 10^{-1}$ | 5.6 days | | | |
| **MS2, 395 b** | NA | | NA | $1.2 (\pm 2.0) \times 10^{-1}$<br>(NS) | $6.8 (\pm 4.6) \times 10^{-1}$ | 6.8 days | | | |
| **BCoV** | $2.2 (\pm 0.61) \times 10^2$ | | 15 minutes | NA | | NA | NA | | NA |
|  | First phase:<br>0-4 hours | Second phase:<br>4-24 hours |  | First phase:<br>0-50 hours | Second phase:<br>50-200 hours |  | First phase:<br>0-50 hours | Second phase:<br>50-400 hours |  |
| **T3** | $3.5 (\pm 1.5) \times 10^1$ | $1.8 (\pm 1.1)$ | 1.6 hours | NA | | NA | NA | | NA |
| **T4** | $3.0 (\pm 0.62) \times 10^1$ | $7.6 (\pm 9.9) \times 10^{-1}$ | 1.9 hours | $3.3 (\pm 2.5) \times 10^{-1}$ | $8.1 (\pm 5.6) \times 10^{-1}$ | 4.1 days | $2.0 (\pm 6.2) \times 10^{-1}$<br>(NS) | $1.0 (\pm 0.41)$ | 3.9 days |

\*NS annotates the rate constants that were not statistically different from zero.

We anticipated that the rapid degradation of the (+)ssRNA was due to the presence of RNase enzymes in wastewater. Indeed, when we repeated the MS2 (+)ssRNA experiment in wastewater samples containing a reagent that inactivates RNase activity (RNAsecure™), the MS2 (+)ssRNA target degraded at a slower rate (Supporting Information Figure S3). Specifically, 0.5-log_10_ decay was observed in the first 30 minutes, and 1.9-log_10_ decay was observed after 4 hours. The role of RNase activity in our untreated wastewater is consistent with previous research on the prevalence of RNases in wastewater.^54^

The extraviral T3 and T4 dsDNA exhibited biphasic kinetics, with faster degradation kinetics over the first 4 hours followed by slower degradation kinetics over the next 20 hours (Figure 1). When modeled as two phases of first-order decay, the resulting rate constants for T3 and T4 are 3.5 (± 1.5) × 10^1^ days^-1^ and 3.0 (± 0.62) × 10^1^ days^-1^ for the first phase (<4 hours), respectively, and 1.8 (± 1.1) days^-1^ and 7.6 (± 9.9) × 10^-1^ days^-1^ for the second phase (>4 hours), respectively. We first hypothesized that the observed slower second phase was due to a small fraction of the nucleic acids that were not effectively extracted from the virus capsids with the nucleic acid extraction kits. However, all the nucleic acids in the dsDNA extracts were susceptible to DNase enzymes and this was not the case for the encapsidated dsDNA (Figure S6); we therefore concluded that the slower kinetics were not due to a small fraction of encapsidated T3 and T4 dsDNA in the extraviral dsDNA stock. We also ruled out the presence of background encapsidated T3 and T4 dsDNA in the wastewater. Specifically, qPCR measurements conducted on the wastewater samples prior to spiking in the viruses did not detect T3 and T4 dsDNA targets at concentrations that would interfere with our experiments (Figure S5). Previous research on environmental DNA (eDNA) in freshwater have reported a protective effect of sediments on DNA persistence.^55,56^ It is therefore possible that the persistent fraction of our DNA was due to extracellular DNA that had partitioned to wastewater solids.

The T_90_ of the extraviral MS2 and BCoV (+)ssRNA was on the order of minutes at 25 °C (Table 2). These results suggest that the viral (+)ssRNA that is quantified in wastewater for wastewater-based epidemiology purposes^1,3,6,18,20^ or for understanding the concentration of pathogens throughout wastewater treatment processes^42,49,57^ is unlikely to be extraviral and that when (+)ssRNA particles are compromised, the RNA genomes will rapidly degrade. The extraviral dsDNA from our two model viruses persisted longer than the extraviral (+)ssRNA, with T_90_ values on the order of 1-2 hours. Given that wastewater spends an average of 3.3 hours in sewershed conveyance, with an upper limit of 36 hours,^33^ it is possible that a small fraction of the viral dsDNA detected in wastewater collected at wastewater treatment plants is from compromised virus particles.

The extraviral (+)ssRNA and dsDNA decay results were highly reproducible in the wastewater samples collected on three different days across an eleven-month period (November to October). This suggests that the nucleic acid decays were not influenced by daily or seasonal variabilities in the wastewater characteristics. Furthermore, the (+)ssRNA of the two different viruses exhibited similar decay kinetics, as did the dsDNA genomes of two different viruses. The two (+)ssRNA viruses and the two dsDNA varied in genome size and structure. Although this is a limited number of nucleic acid molecules and wastewater from a single wastewater treatment plant, the results suggest that different extraviral (+)ssRNA and dsDNA have similar degradation kinetics in wastewater.

### Encapsidated RNA and DNA degradation kinetics

The encapsidated MS2 (+)ssRNA and T4 dsDNA degraded with biphasic kinetics (Figure 2) and the overall degradation was slower than the extraviral (+)ssRNA and dsDNA degradation (Table 2). Whereas the extraviral nucleic acid decay kinetics slowed over time (Figure 1), the kinetics of the encapsidated (+)ssRNA and dsDNA accelerated over time (Figure 2). The MS2 (+)ssRNA target concentration was nearly stable for the first 2-3 days in wastewater, and then began to decrease after 3-4 days. The dsDNA kinetics began to accelerate after the first 2 days. When modeled as two phases of first-order kinetics, the initial first phase rate constants were on the order of 0.1-0.3 days^-1^ and the second phase rate constants on the order of 0.6-0.8 days^-1^ for MS2 and T4 (Figure 2; Table 2). Both the encapsidated MS2 (+)ssRNA and T4 dsDNA underwent little decay in the PBS control samples (Figure S7). Combined, the extraviral and encapsidated viral genome results demonstrate the protective effect of viral capsids on (+)ssRNA of MS2 and the dsDNA of T4 in municipal wastewater.

The T_90_ for the encapsidated (+)ssRNA was 5.6 days and the T_90_ for the dsDNA was 4.1 days (Table 2). These results suggest that the viral nucleic acids of intact virus particles are stable over the time period that most human viruses spend in sewer systems (Table 2). Our experiments were conducted at 25 °C, and virus nucleic acid signals degrade faster in wastewater at warmer temperatures.^58–60^ The encapsidated MS2 (+)ssRNA kinetics measured here at 25 °C are comparable to kinetics reported for several other (+)ssRNA viruses (Table S6). In those studies, the first-order decay rate constants for (+)ssRNA viruses (both naked and enveloped) ranged from 0.09-0.84 days^-1^ and the T_90_ values ranged from 2.74-26 days. Other studies observed biphasic kinetics of (+)ssRNA decay in wastewater, including for SARS-CoV-2 and MHV (+)ssRNA.^59,61^ Autoclaved wastewater led to longer persistence of SARS-CoV-2 and MHV (+)ssRNA likely due to decreases in biological activity.^58^ To our knowledge, viral dsDNA decay kinetics in municipal wastewater have not been previously reported. Our data, combined with decay rates from literature, suggest that (+)ssRNA and dsDNA susceptibility is not the primary determinant of viral nucleic acid detection in wastewater.

### Effect of amplicon size on genome persistence

We hypothesized that the assay amplicon size would affect the observed persistence of the encapsidated nucleic acids, with longer targets degrading faster than shorter targets. This effect has been observed when nucleic acids react with disinfectants, such as free chlorine and UV_254_.^25,43^ To test if this was true for viral genome degradation in wastewater, we conducted wastewater experiments with 3 sets of molecular assays that targeted the same region of the MS2 (+)ssRNA genome and that had increasing amplicon sizes (99 b, 192 b, 395 b). When the MS2 virus was incubated at 25 °C in untreated wastewater, the different amplicon targets degraded with similar biphasic kinetics (Figure 3) and there was no statistical difference between the decay kinetics of the three amplicons in each of the two modeled kinetic phases. We anticipate this is because the MS2 genome begins to degrade only after the virus particles are compromised and that once compromised, the rapid decay of extraviral (+)ssRNA leads to similar observed decay kinetics for the three amplicon sizes. This finding that (+)ssRNA amplicon sizes between 99 and 395 bases have the same degradation kinetics in wastewater is important for WBE, as it indicates that the absolute values quantified in wastewater may not be impacted by assays that target different amplicon sizes.

**Figure 3.**
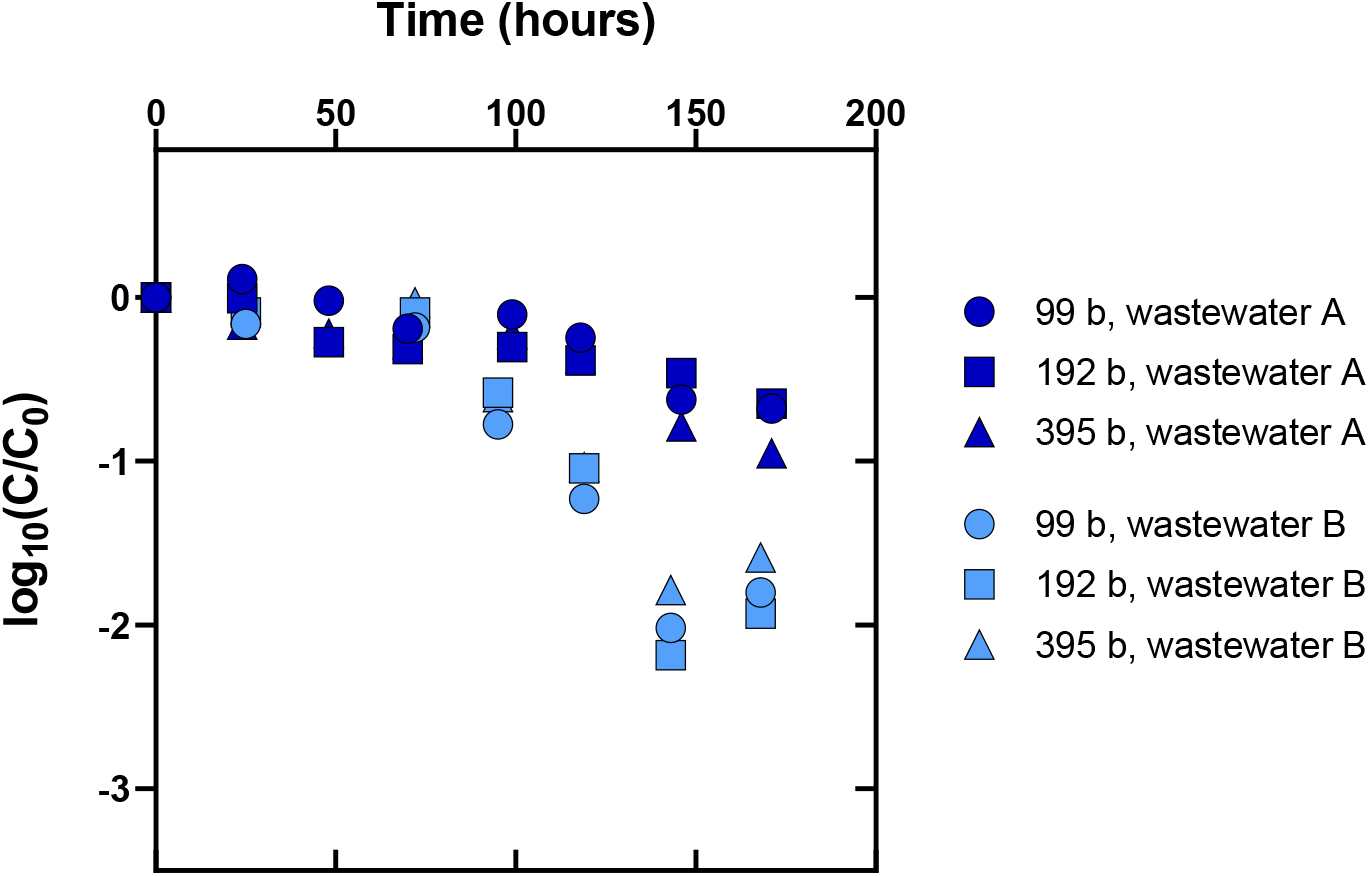
Effect of three different amplicon sizes on encapsidated MS2 genome persistence in untreated wastewater at 25 °C. C refers to the concentration of the viral nucleic acids in gene copies μL^-1^ at time t in hours. C_0_ represents the initial concentration of the viral nucleic acids at time = 0 based on the amount of gene copies measured immediately after the stocks were added to the sample. Experiments were run in duplicate with two different wastewater samples collected on different days.

### Comparison of infectivity and nucleic acid degradation

We measured the loss of the (+)ssRNA MS2 and dsDNA T4 infectivity in the same wastewater samples that we measured the encapsidated nucleic acid decay kinetics (Figure 4). The inactivation kinetics of the MS2 and T4 viruses varied between experimental replicates and the kinetics were biphasic, with an initial lag phase followed by an acceleration. The mean T_90_ values that incorporated both phases equaled 4.3 days and 3.9 days for MS2 and T4, respectively (Table 2). The inactivation kinetics varied between experimental replicates, with T_90_ values varying between 3.3 and 5.5 days for MS2 and 2.5 and 4.4 days for T4 for the three different wastewater samples. A previous study by our laboratory reported a T_90_ value of 5.0 days for MS2^49^ and T_90_ values for enveloped virus MHV and enveloped bacteriophage *ϕ*6 as 0.5 days and 0.3 days, respectively.^49^ Other labs have reported T_90_ values of ∼2 days for enveloped SARS-CoV-2^61^ and 3-6 days for naked Poliovirus types 2 and 3.^62^ We note that the enveloped SARS-CoV-2 virus is understood to be mostly non-infectious when it enters wastewater.

**Figure 4.**
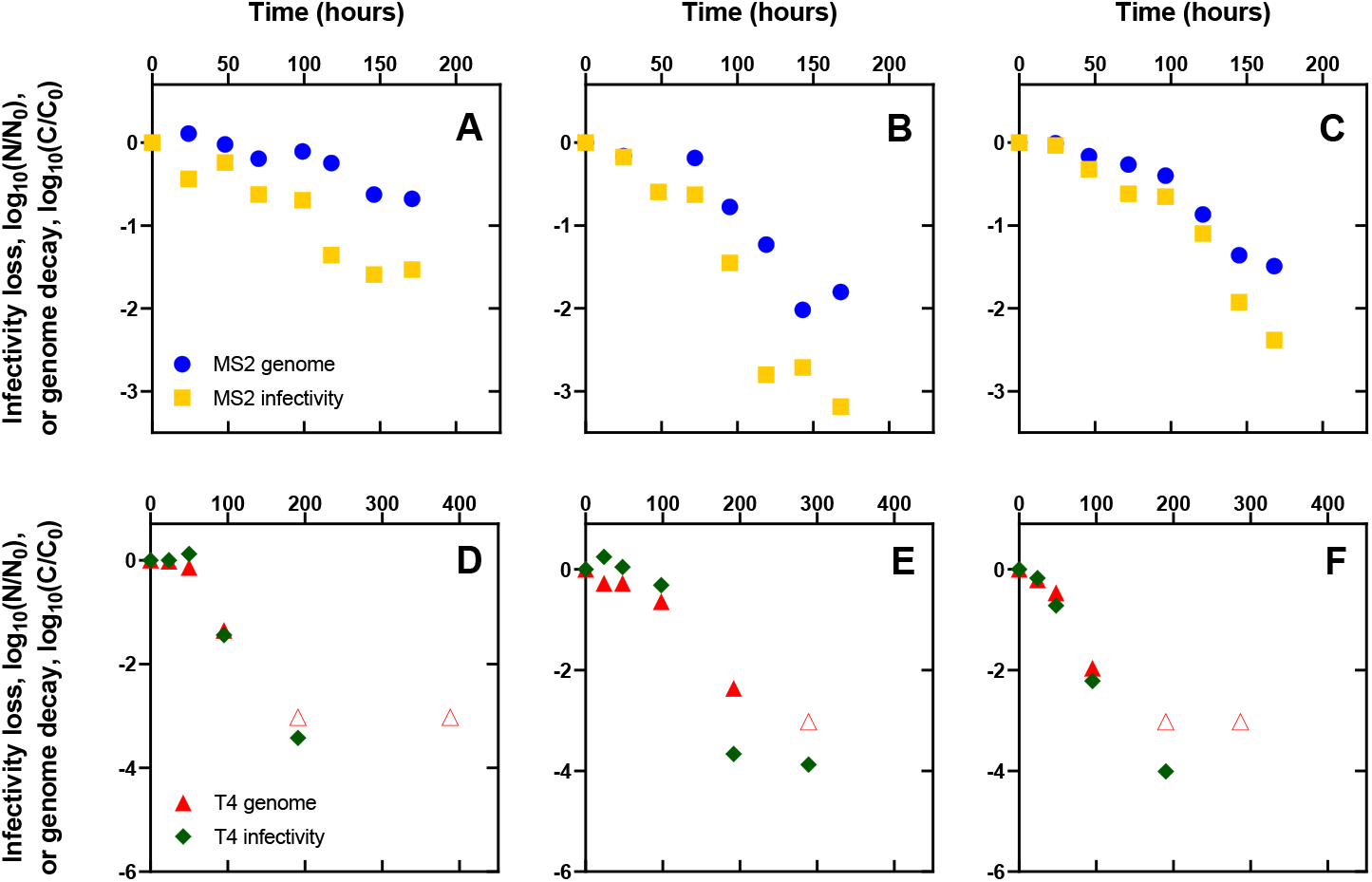
Infectivity loss and genome decay for MS2 (panels A, B, C) and T4 (panels D, E, F) in untreated wastewater at 25 °C. C refers to the concentration of the viral nucleic acids in gene copies μL^-1^ at time t in hours. C_0_ represents the initial concentration of the viral nucleic acids at time = 0 based on the amount of gene copies measured immediately after the stocks were added to the sample. N refers to the concentration of infectious viruses in PFU mL^-1^ at time t in hours. N_0_ represents the initial concentration of the infectious virus at time = 0 based on the number of plaques measured immediately after the stocks were added to the sample. Open symbols represent measurements that were below the limit of quantification and are plotted at the limit of quantification for each virus assay. We present the triplicate experimental replicates for each virus in three different wastewater samples in different panels for clarity.

In the individual experiments, the inactivation kinetics of infectious T4 nearly matched the respective dsDNA genome signal decay, whereas the MS2 (+)ssRNA decay followed shortly behind the loss in infectivity (Figure 4). We also observed very little difference between first-order decay rates for encapsidated (+)ssRNA viral nucleic acids and encapsidated dsDNA viral nucleic acids in untreated wastewater at 25 °C (Table 2). These results with MS2 (+)ssRNA and T4 dsDNA in wastewater at room temperature suggest that the nucleic acids break down very soon after the viruses are inactivated and that protein capsid characteristics, rather than genome type, controls the virus nucleic acid signal in wastewater. Previously, Kline et al. observed a T_90_ of 3 days for naked Poliovirus type 3 infectivity in wastewater at room temperature, whereas the molecular signal measured by RT-qPCR had a T_90_ of 7 days.^62^ In surface waters at room temperature, the difference between the infectivity kinetics and genome kinetics is much more pronounced, with the naked norovirus genome signal lasting much longer than the norovirus infectivity.^26^

The observed differences in the inactivation rates between viruses, and the relative stability of the infectious virus and the nucleic acids are likely a result of the differences in the microbiology of the different waters and mechanisms of virus inactivation in the distinct waters. Municipal wastewater contains a wide range of bacteria and protozoa, both of which have been linked with virus inactivation.^38,39^ In lake water, bacterial proteases were responsible for most of the observed inactivation of enteroviruses E11 and CVA9 and the two viruses had different susceptibilities to the bacterial proteases.^39^ Furthermore, an earlier study demonstrated that the exposure of Coxsackievirus type A9 to *P. aeruginosa* proteases lead to the inactivation and subsequent release of viral (+)ssRNA.^63^ Based on our results combined with literature on bacterial proteases in surface waters, we hypothesize that proteases play a significant role in naked virus inactivation in wastewater and that this mechanism leads to the release of nucleic acids. Once the nucleic acids are released into the wastewater, they are rapidly degraded by nucleases native to wastewater. The different decay rates of encapsidated nucleic acids in wastewater observed in this study and between studies is likely a combination of varying protease activity in wastewater samples as well as the different susceptibilities of the viruses to wastewater proteases.

## ENVIRONMENTAL IMPLICATIONS

In this study, we report rate constants for model (+)ssRNA and dsDNA, both extraviral and encapsidated, in wastewater at 25 °C and compare nucleic acid decay with virus inactivation. The decay rates for extraviral nucleic acids were comparable for the two (+)ssRNA genomes and also for the two dsDNA viral genomes. Decay rates were also consistent in wastewater samples that were collected across multiple seasons from a single wastewater treatment plant. Our results show that extraviral nucleic acids that are excreted into the wastewater or released from microorganisms as wastewater is transported to treatment plants are degraded within minutes for (+)ssRNA or hours for dsDNA. A small fraction (∼0.1%) of the spiked extraviral dsDNA targets was persistent for greater than 24 hours, possibly due to protection from wastewater solids. Future work should investigate the impact of wastewater solids partitioning on nucleic acid stability. Overall, these results suggest that the viral (+)ssRNA and dsDNA signals measured at the wastewater treatment plant are predominantly part of intact virus particles. These findings have implications for recovery method development and optimization since the relative contribution of extraviral nucleic acids are unlikely to have a large effect on the total quantities of virus gene copy numbers.

The virus capsids of MS2 and T4 provided major protection to the nucleic acid signals. At 25 °C, the T_90_ values for the MS2 (+)ssRNA and T4 dsDNA genome targets increased from 19 minutes and 1.9 hours when extraviral to 5.6 days and 4.1 days when encapsidated, respectively. Our results suggest that capsid stability is the primary determinant of genome stability, and thus detection, in wastewater. The slow decay kinetics of our model virus encapsidated genomes confirm that the encapsidated nucleic acid signals are effectively stable over the period of times it takes for wastewater to be conveyed to wastewater treatment plants.

The virus genome signals degraded soon after the viruses lost infectivity and, in some replicates, appeared to occur concurrently. These findings are relevant for when gene copy measurements conducted in wastewater are used to determine human health risks from enteric viruses. Human norovirus, for example, is difficult to culture *in vitro* and therefore direct potable reuse crediting frameworks rely on molecular measurements to determine how much credit is given to a particular water treatment process.^24^ If these results for MS2 and T4 are representative of naked human viruses, then it follows that the infectious virus to gene copy ratios measured at the wastewater treatment plant are likely similar to the ratios originally excreted. Bacteriophage MS2 and T4 are naked (i.e. non-enveloped) viruses, and the relationship between gene copies and infectivity may be different for enveloped viruses due to their increased inactivation rate constants in wastewater.^49,61^ Although more research should be conducted for different viruses, wastewaters, and temperatures, these results strengthen the case for using enteric virus molecular signals in wastewater as an indicator of infectious virus levels that may impact human health.

Finally, we show that the amplicon sizes over the range of 99-395 bases exhibited similar degradation kinetics in untreated wastewater at room temperature for one region of the MS2 genome. This suggests that various amplicon sizes employed in wastewater (+)ssRNA virus measurements do not have a major effect on the quantities of gene copies measured. Ultimately, (+)ssRNA virus quantities that are measured with different assays that target different amplicon sizes should be directly comparable.

We note that our experiments were limited to wastewater at 25 °C and wastewater temperature has a pronounced effect on virus inactivation kinetics and viral genome stability.^58–60^ Future work will need to confirm that the trends observed here between extraviral (+)ssRNA and extraviral dsDNA, and between encapsidated and extraviral nucleic acids, are maintained at different temperatures. The group of model viruses was limited, and an expanded group of viruses, as well as dsRNA and ssDNA genome types, should be studied in the future. Despite these limitations, we anticipate our results will aid the interpretation and application of virus wastewater measurements, both for wastewater epidemiology and microbial risk assessment applications.

## Data Availability

All data produced in the present study are available upon reasonable request to the authors.

## References

1. Graham, K. E. et al. SARS-CoV-2 RNA in Wastewater Settled Solids Is Associated with COVID-19 Cases in a Large Urban Sewershed. Environ. Sci. Technol. 55, 488–498 (2021).

2. Ahmed, W. et al. First confirmed detection of SARS-CoV-2 in untreated wastewater in Australia: A proof of concept for the wastewater surveillance of COVID-19 in the community. Sci. Total Environ. 728, 138764 (2020).

3. Soller, J. et al. Modeling infection from SARS-CoV-2 wastewater concentrations: promise, limitations, and future directions. J. Water Health 20, 1197–1211 (2022).

4. Aziz, M. A. et al. Environmental surveillance of SARS-CoV-2 in municipal wastewater to monitor COVID-19 status in urban clusters in Malaysia. Arch. Microbiol. 205, 76 (2023).

5. Medema, G., Heijnen, L., Elsinga, G., Italiaander, R. & Brouwer, A. Presence of SARS-Coronavirus-2 RNA in Sewage and Correlation with Reported COVID-19 Prevalence in the Early Stage of the Epidemic in The Netherlands. Environ. Sci. Technol. Lett. 7, 511–516 (2020).

6. Gonzalez, R. et al. COVID-19 surveillance in Southeastern Virginia using wastewater-based epidemiology. Water Res. 186, 116296 (2020).

7. Westhaus, S. et al. Detection of SARS-CoV-2 in raw and treated wastewater in Germany – Suitability for COVID-19 surveillance and potential transmission risks. Sci. Total Environ. 751, 141750 (2021).

8. Tanhaei, M. et al. The first detection of SARS-CoV-2 RNA in the wastewater of Tehran, Iran. Environ. Sci. Pollut. Res. 28, 38629–38636 (2021).

9. Haramoto, E., Malla, B., Thakali, O. & Kitajima, M. First environmental surveillance for the presence of SARS-CoV-2 RNA in wastewater and river water in Japan. Sci. Total Environ. 737, 140405 (2020).

10. Hata, A., Hara-Yamamura, H., Meuchi, Y., Imai, S. & Honda, R. Detection of SARS-CoV-2 in wastewater in Japan during a COVID-19 outbreak. Sci. Total Environ. 758, 143578 (2021).

11. Hasan, S. W. et al. Detection and quantification of SARS-CoV-2 RNA in wastewater and treated effluents: Surveillance of COVID-19 epidemic in the United Arab Emirates. Sci. Total Environ. 764, 142929 (2021).

12. Chakraborty, P. et al. First surveillance of SARS-CoV-2 and organic tracers in community wastewater during post lockdown in Chennai, South India: Methods, occurrence and concurrence. Sci. Total Environ. 778, 146252 (2021).

13. Kumar, M. et al. First proof of the capability of wastewater surveillance for COVID-19 in India through detection of genetic material of SARS-CoV-2. Sci. Total Environ. 746, 141326 (2020).

14. Wolken, M. et al. Wastewater surveillance of SARS-CoV-2 and influenza in preK-12 schools shows school, community, and citywide infections. Water Res. 231, 119648 (2023).

15. Miyani, B., Fonoll, X., Norton, J., Mehrotra, A. & Xagoraraki, I. SARS-CoV-2 in Detroit Wastewater. J. Environ. Eng. 146, 06020004 (2020).

16. Lopez Marin, M. A. et al. Monitoring COVID-19 spread in selected Prague’s schools based on the presence of SARS-CoV-2 RNA in wastewater. Sci. Total Environ. 871, 161935 (2023).

17. Aschidamini Prandi, B. et al. Long Term Wastewater Based Epidemiology Of Sars-Cov-2 And Social Restrictions Applied In Porto Alegre, Southern Brazil. Sci. One Health 100008 (2023) doi:10.1016/j.soh.2023.100008.

18. Wolfe, M. K. et al. Wastewater-Based Detection of Two Influenza Outbreaks. Environ. Sci. Technol. Lett. 9, 687–692 (2022).

19. Wolfe, M. K. et al. Wastewater Surveillance for Monkeypox Virus in Nine California Communities. 2022.09.06.22279312 Preprint at https://doi.org/10.1101/2022.09.06.22279312 (2022).

20. Hughes, B. et al. Respiratory Syncytial Virus (RSV) RNA in Wastewater Settled Solids Reflects RSV Clinical Positivity Rates. Environ. Sci. Technol. Lett. 9, 173–178 (2022).

21. Kazama, S. et al. Environmental Surveillance of Norovirus Genogroups I and II for Sensitive Detection of Epidemic Variants. Appl. Environ. Microbiol. 83, e03406–16 (2017).

22. Vo, V. et al. Identification and genome sequencing of an influenza H3N2 variant in wastewater from elementary schools during a surge of influenza A cases in Las Vegas, Nevada. Sci. Total Environ. 162058 (2023) doi:10.1016/j.scitotenv.2023.162058.

23. Rockey, N. et al. UV Disinfection of Human Norovirus: Evaluating Infectivity Using a Genome-Wide PCR-Based Approach. Environ. Sci. Technol. 54, 2851–2858 (2020).

24. Jahne, M. A. et al. Enteric pathogen reduction targets for onsite non-potable water systems: A critical evaluation. Water Res. 233, 119742 (2023).

25. Pecson, B. M., Ackermann, M. & Kohn, T. Framework for Using Quantitative PCR as a Nonculture Based Method To Estimate Virus Infectivity. Environ. Sci. Technol. 45, 2257–2263 (2011).

26. Shaffer, M., Huynh, K., Costantini, V., Bibby, K. & Vinjé, J. Viable Norovirus Persistence in Water Microcosms. Environ. Sci. Technol. Lett. 9, 851–855 (2022).

27. Hodges, K. & Gill, R. Infectious diarrhea. Gut Microbes 1, 4–21 (2010).

28. Zhang, Y. et al. Prevalence and Persistent Shedding of Fecal SARS-CoV-2 RNA in Patients With COVID-19 Infection: A Systematic Review and Meta-analysis. Clin. Transl. Gastroenterol. 12, e00343 (2021).

29. Zang, R. et al. TMPRSS2 and TMPRSS4 promote SARS-CoV-2 infection of human small intestinal enterocytes. Sci. Immunol. 5, eabc3582 (2020).

30. Long, S. SARS-CoV-2 Subgenomic RNAs: Characterization, Utility, and Perspectives. Viruses 13, 1923 (2021).

31. Miller, W. A. & Koev, G. Synthesis of Subgenomic RNAs by Positive-Strand RNA Viruses. Virology 273, 1–8 (2000).

32. Alexandersen, S., Chamings, A. & Bhatta, T. R. SARS-CoV-2 genomic and subgenomic RNAs in diagnostic samples are not an indicator of active replication. Nat. Commun. 11, 6059 (2020).

33. Kapo, K. E., Paschka, M., Vamshi, R., Sebasky, M. & McDonough, K. Estimation of U.S. sewer residence time distributions for national-scale risk assessment of down-the-drain chemicals. Sci. Total Environ. 603–604, 445–452 (2017).

34. McCall, C. et al. Modeling SARS-CoV-2 RNA degradation in small and large sewersheds. Environ. Sci. Water Res. Technol. 8, 290–300 (2022).

35. Burnet, J.-B., Cauchie, H.-M., Walczak, C., Goeders, N. & Ogorzaly, L. Persistence of endogenous RNA biomarkers of SARS-CoV-2 and PMMoV in raw wastewater: Impact of temperature and implications for wastewater-based epidemiology. Sci. Total Environ. 857, 159401 (2023).

36. Guo, Y., Sivakumar, M. & Jiang, G. Decay of four enteric pathogens and implications to wastewater-based epidemiology: Effects of temperature and wastewater dilutions. Sci. Total Environ. 819, 152000 (2022).

37. Chattopadhyay, D., Chattopadhyay, S., Lyon, W. G. & Wilson, J. T. Effect of Surfactants on the Survival and Sorption of Viruses. Environ. Sci. Technol. 36, 4017–4024 (2002).

38. Olive, M., Gan, C., Carratalà, A. & Kohn, T. Control of Waterborne Human Viruses by Indigenous Bacteria and Protists Is Influenced by Temperature, Virus Type, and Microbial Species. Appl. Environ. Microbiol. 86, e01992–19 (2020).

39. Corre, M.-H., Bachmann, V. & Kohn, T. Bacterial matrix metalloproteases and serine proteases contribute to the extra-host inactivation of enteroviruses in lake water. ISME J. 16, 1970–1979 (2022).

40. Wolfe, M. K. et al. Scaling of SARS-CoV-2 RNA in Settled Solids from Multiple Wastewater Treatment Plants to Compare Incidence Rates of Laboratory-Confirmed COVID-19 in Their Sewersheds. Environ. Sci. Technol. Lett. 8, 398–404 (2021).

41. Wolfe, M. et al. Detection of SARS-CoV-2 Variants Mu, Beta, Gamma, Lambda, Delta, Alpha, and Omicron in Wastewater Settled Solids Using Mutation-Specific Assays Is Associated with Regional Detection of Variants in Clinical Samples. Appl. Environ. Microbiol. 88, e00045–22 (2022).

42. Qiao, Z. et al. Reactivity of Viral Nucleic Acids with Chlorine and the Impact of Virus Encapsidation. Environ. Sci. Technol. 56, 218–227 (2022).

43. He, H. et al. Degradation and Deactivation of Bacterial Antibiotic Resistance Genes during Exposure to Free Chlorine, Monochloramine, Chlorine Dioxide, Ozone, Ultraviolet Light, and Hydroxyl Radical. Environ. Sci. Technol. 53, 2013–2026 (2019).

44. Jo, T. et al. Rapid degradation of longer DNA fragments enables the improved estimation of distribution and biomass using environmental DNA. Mol. Ecol. Resour. 17, e25–e33 (2017).

45. Mesquita, M. M. F. et al. Optimal preparation and purification of PRD1-like bacteriophages for use in environmental fate and transport studies. Water Res. 44, 1114–1125 (2010).

46. Adams, M. H. Bacteriophages. Bacteriophages. (1959).

47. Pecson, B. M., Martin, L. V. & Kohn, T. Quantitative PCR for determining the infectivity of bacteriophage MS2 upon inactivation by heat, UV-B radiation, and singlet oxygen: advantages and limitations of an enzymatic treatment to reduce false-positive results. Appl. Environ. Microbiol. 75, 5544–5554 (2009).

48. Kantor, R. S., Nelson, K. L., Greenwald, H. D. & Kennedy, L. C. Challenges in Measuring the Recovery of SARS-CoV-2 from Wastewater. Environ. Sci. Technol. 55, 3514–3519 (2021).

49. Ye, Y., Ellenberg, R. M., Graham, K. E. & Wigginton, K. R. Survivability, Partitioning, and Recovery of Enveloped Viruses in Untreated Municipal Wastewater. Environ. Sci. Technol. 50, 5077–5085 (2016).

50. Borchardt, M. A. et al. The Environmental Microbiology Minimum Information (EMMI) Guidelines: qPCR and dPCR Quality and Reporting for Environmental Microbiology. Environ. Sci. Technol. 55, 10210–10223 (2021).

51. Decaro, N. et al. Detection of bovine coronavirus using a TaqMan-based real-time RT-PCR assay. J. Virol. Methods 151, 167–171 (2008).

52. Kettleson, E. M. et al. Airborne Virus Capture and Inactivation by an Electrostatic Particle Collector. Environ. Sci. Technol. 43, 5940–5946 (2009).

53. Lim, S. W., Lance, S. T., Stedman, K. M. & Abate, A. R. PCR-activated cell sorting as a general, cultivation-free method for high-throughput identification and enrichment of virus hosts. J. Virol. Methods 242, 14–21 (2017).

54. Holm, R. H. et al. Surveillance of RNase P, PMMoV, and CrAssphage in wastewater as indicators of human fecal concentration across urban sewer neighborhoods, Kentucky. FEMS Microbes 3, xtac003 (2022).

55. Sakata, M. K. et al. Sedimentary eDNA provides different information on timescale and fish species composition compared with aqueous eDNA. Environ. DNA 2, 505–518 (2020).

56. Barnes, M. A. et al. Environmental Conditions Influence eDNA Persistence in Aquatic Systems. Environ. Sci. Technol. 48, 1819–1827 (2014).

57. Ye, Y., Chang, P. H., Hartert, J. & Wigginton, K. R. Reactivity of Enveloped Virus Genome, Proteins, and Lipids with Free Chlorine and UV 254. Environ. Sci. Technol. 52, 7698–7708 (2018).

58. Ahmed, W. et al. Decay of SARS-CoV-2 and surrogate murine hepatitis virus RNA in untreated wastewater to inform application in wastewater-based epidemiology. Environ. Res. 191, 110092 (2020).

59. Chandra, F. et al. Persistence of Dengue (Serotypes 2 and 3), Zika, Yellow Fever, and Murine Hepatitis Virus RNA in Untreated Wastewater. Environ. Sci. Technol. Lett. 8, 785–791 (2021).

60. Muirhead, A. et al. Zika Virus RNA Persistence in Sewage. Environ. Sci. Technol. Lett. 7, 659–664 (2020).

61. Bivins, A. et al. Persistence of SARS-CoV-2 in Water and Wastewater. Environ. Sci. Technol. Lett. 7, 937–942 (2020).

62. Kline, A. et al. Persistence of poliovirus types 2 and 3 in waste-impacted water and sediment. PLOS ONE 17, e0262761 (2022).

63. Cliver, D. O. & Herrmann, J. E. Proteolytic and microbial inactivation of enteroviruses. Water Res. 6, 797–805 (1972).

64. Hart, O. E. & Halden, R. U. Modeling wastewater temperature and attenuation of sewage-borne biomarkers globally. Water Res. 172, 115473 (2020).

