## Supporting Information for "The Protective Effect of Virus Capsids on RNA and DNA Virus Genomes in Wastewater"

Number of Pages: 23

Number of Tables: 6

Number of Figures: 8

### Table of Contents


***Table S1.*** *Summary of media recipes and growth conditions for bacteriophage* ...... **S3**

***Table S2.*** *Variation in pH between matrices used during this study* ...... **S4**

***Table S3.*** *Variation in solids of 24-hour composite raw influent samples during this study* ...... **S4**

***Example $T_{90}$ calculation for two phases*** ...... **S5**

***RNase treatment*** ...... **S6**

***Figure S1.*** *MS2 gene copy concentrations of RNase treated and untreated MS2 stock used for experiments* ...... **S7**

***DNase treatment*** ...... **S8**

***Figure S2.*** *T4 gene copy concentrations of DNase treated and untreated T4 stock used for experiments* ...... **S9**

***RNase-free experiments*** ...... **S10**

***Figure S3.*** *MS2 ssRNA persistence in untreated wastewater with the addition of RNASecure at 25℃* .. **S11**

***qPCR assays*** ...... **S12**

***Table S4.*** *qPCR assays for virus targets* ...... **S14**

***Figure S4.*** *Comparison of estimated gene copies from diluted sample with actual gene copies in samples* ...... **S15**

***Figure S5.*** *Measured virus targets in wastewater background* ...... **S16**

***ddPCR assays*** ...... **S17**

***Table S5.*** *ddPCR assays for virus targets* ...... **S18**

***Figure S6.*** *DNase treatment of T3 and T4 genomes to ensure extraviral nucleic acids* ...... **S19**

***Table S6.*** *Summary of reported decay rates for encapsidated viral nucleic acids in wastewater* ...... **S20**

***Figure S7.*** *Extraviral (+)ssRNA and dsDNA decay in PBS at 25℃* ...... **S21**

***Figure S8.*** *Encapsidated MS2 (+)ssRNA and T4 dsDNA genome decay in PBS at 25℃* ...... **S22**

***References*** ...... **S23**

***Table S1.*** *Summary of media recipes and growth conditions for bacteriophage*

| Phage | Bacterial Host | Media Broth (1L) | 0.7% Soft Agar (1L) | 1.5% Hard Agar (1L) | Incubation Temp/Time |
|---|---|---|---|---|---|
| MS2 | *E. coli* ATCC 15597 | 10.0g Tryptone (BD 211705)<br><br>1.0g Yeast extract<br><br>8.0g NaCl<br><br>1.0g D-glucose<br><br>0.22g CaCl2<br><br>10mL Thiamine (1mg/mL) | 10.0g Tryptone (BD 211705)<br><br>1.0g Yeast extract<br><br>8.0g NaCl<br><br>1.0g D-glucose<br><br>0.22g CaCl2<br><br>10mL Thiamine (1mg/mL)<br><br>*7.0g Agar* | 10.0g Tryptone (BD 211705)<br><br>1.0g Yeast extract<br><br>8.0g NaCl<br><br>1.0g D-glucose<br><br>0.22g CaCl2<br><br>10mL Thiamine (1mg/mL)<br><br>*15.0g Agar* | 37$^o$C, overnight (12+ hours) |
| T3/T4 | *E. coli* ATCC 11303 | 8.0g Nutrient broth (BD 234000)<br><br>5.0g NaCl | 8.0g Nutrient broth (BD 234000)<br><br>5.0g NaCl<br><br>*7.0g Agar* | 8.0g Nutrient broth (BD 234000)<br><br>5.0g NaCl<br><br>*15.0g Agar* | T3: 37$^o$C, *~5-6 hours*<br><br>T4: 37$^o$C, overnight (12+ hours) |

***Table S2.*** *Variation in pH between matrices used during this study*

| Sample Type | Mean pH (min to max) | Mean change in pH during experiments |
|---|---|---|
| Wastewater influent | 7.74<br>(7.32 to 8.00) | 0.95 |
| 10X PBS | 7.43<br>(7.34 to 7.48) | 0.03 |
| 1X PBS | 7.86<br>(7.47 to 7.99) | 0.03 |

***Table S3.*** *Variation in solids of 24-hour composite raw influent samples during this study*

| Parameter | Mean value (min to max) | Standard deviation |
|---|---|---|
| TSS | 196<br>(68 to 312) | 51.4 |
| TVSS | 166<br>(64 to 216) | 36.5 |

***Example $T_{90}$ calculation for two phases.*** In cases where biphasic kinetics were observed, the two phases were modeled separately by linear regression. The $T_{90}$ value was determined by the decay in the first phase and the decay in the second phase that added to 1-$\log_{10}$ removal of the virus. This was calculated for *T4 inactivation kinetics* as follows:

First phase: 0-50 hours

Second phase: 50-400 hours

First phase $k_1$ = 0.003631 (± 0.011139) hours$^{-1}$

Second phase $k_2$ = 0.01869 (± 0.00744) hours$^{-1}$

Inactivation in the first phase (0-50 hours) = 0.003631 hours$^{-1}$ × 50 hours = 0.18155 $\log_{10}$

Since 0.18155 < 1, we need to compute how many more hours in the second phase of kinetics before the inactivation reaches 1-$\log_{10}$.

Inactivation = 1 – 0.18155 = 0.81845

Hours in the second phase = 0.81845/0.01869 = 43.7908 hours

Total $T_{90}$ from first and second phase = 50 + 43.7908 = 93.7908 hours = 3.9 days

***RNase treatment.*** In order to ensure that most (> 99%) of the MS2 bacteriophage stock was initially encapsidated at the start of the experiments and that extraviral RNA was not a component in the initial stock, an RNase treatment of the stock was performed using a previously established method.[1] Briefly, 20 units of RNase ONE Ribonuclease (10 U $\mu L^{-1}$) (Promega) were added to final sample volumes of 220 µL while untreated controls consisted of 220 µL of sample without RNase ONE. All samples were shaken and incubated at 37°C for 15 minutes. Next, 80 units of RNasin Inhibitor (40 U $\mu L^{-1}$) (Promega) were added to stop the RNase reaction. Samples were incubated at 25°C for 15 minutes without shaking. Untreated and RNase treated samples were run in parallel and in triplicate. Finally, RNA was extracted as described in the RNA and DNA extraction methods section and gene copies were enumerated using ddPCR. A summary figure comparing MS2 gene copies $mL^{-1}$ in the untreated and RNase treated samples is shown in Figure S1. Individual data points represent separate samples run in triplicate (n = 3). For untreated samples, two dilutions ($10^4$ and $10^5$) from triplicate samples were acceptable for ddPCR and included in analysis (n = 6). The black horizontal bar represents the average MS2 gene copies $mL^{-1}$ of the data points in each category.

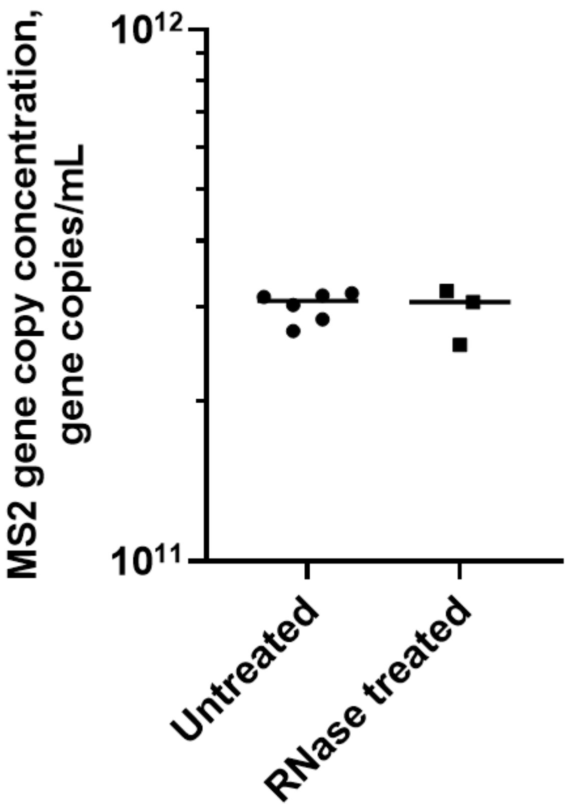


***Figure S1.*** *MS2 gene copy concentrations of RNase treated and untreated MS2 stock used for experiments. Lines depict the mean of six (untreated) or three (RNase treated) independent replicates (n = 6, n = 3).*

***DNase treatment.*** In order to ensure that most (> 99%) of the T4 bacteriophage stock was initially encapsidated at the start of the experiments and that extraviral DNA was not a component in the initial stock, a DNase treatment of the stock was performed using a previously established method.[2] Two buffers were prepared for the DNase treatment. A storage buffer contained 10mM Tris-Cl (pH 7.5), and 2 mM $CaCl_2$ in 50% glycerol. A 10x reaction buffer contained 100 mM Tris-HCl (pH 7.6), 25 mM $MgCl_2$, and 5 mM $CaCl_2$. DNase I (Roche, Cat. No. 10104159001) was added to the in storage buffer at a concentration of 40,000 U $mL^{-1}$ and stored at -20°C. Prior to DNase treatment of samples, DNase I in storage buffer (40,000 U $mL^{-1}$) was diluted 1:40 in the 10x reaction buffer to a concentration of 1,000 U $mL^{-1}$. Stock samples were treated with a final concentration of 100 U $mL^{-1}$ DNase I in 10x reaction buffer for one hour at 25°C without shaking. Untreated samples were not treated with DNase I and were incubated for one hour at 25°C without shaking. Untreated and DNase treated samples were run in parallel and in triplicate. After one hour, 100 mM EDTA and 100 mM EGTA were added to stop the DNase reaction. Finally, DNA was extracted as described in the RNA and DNA extraction methods section and gene copies were enumerated using ddPCR. A summary figure comparing T4 gene copies $mL^{-1}$ in the untreated and DNase treated samples is shown in Figure S2. Individual data points represent individual samples run in triplicate (n = 3). The black horizontal bar represents the average T4 gene copies $mL^{-1}$ of the data points in each category.

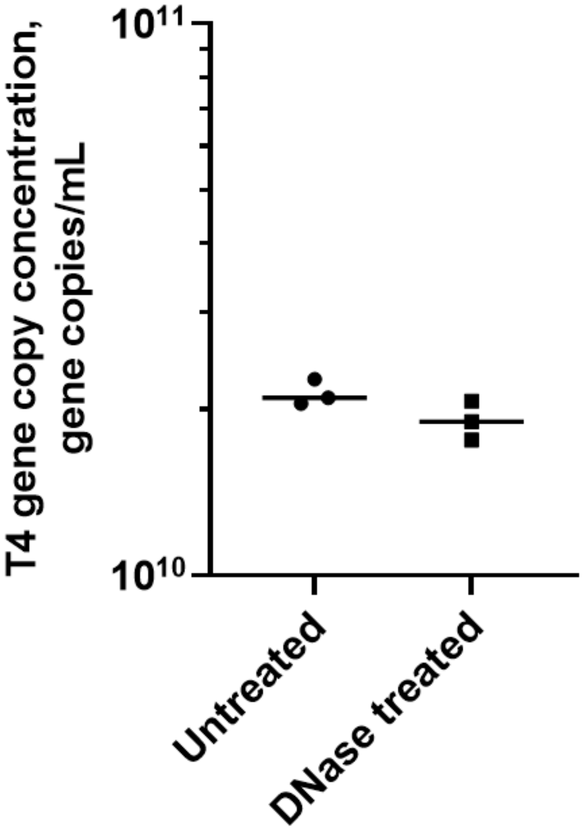


***Figure S2.*** *T4 gene copy concentrations of DNase treated and untreated T4 stock used for experiments. Lines depict the mean of three independent replicates (n = 3).*

***RNase-free experiments.*** Due to the high concentration of RNase enzymes present in wastewater, experiments were designed to inactivate inherent RNases and measure RNA decay in the absence of RNase enzymes. Invitrogen™ RNAsecure™ RNase Inactivation Reagent (Fisher, Cat. No. AM7006) was used to inactivate RNases naturally present in the wastewater matrices. Briefly, RNAsecure™ was added to untreated wastewater, filtered wastewater, and PBS at 1X final concentration. Controls were run in parallel without the addition of the RNAsecure™ reagent. All samples were then heated at 60℃ for 10 minutes according to manufacturer's instructions to inactivate RNases. Heating was performed using an Eppendorf ThermoStat plus microplate heating block. Extraviral RNA was then added to samples for experimental analysis of RNA decay in the presence and absence of RNase enzymes.

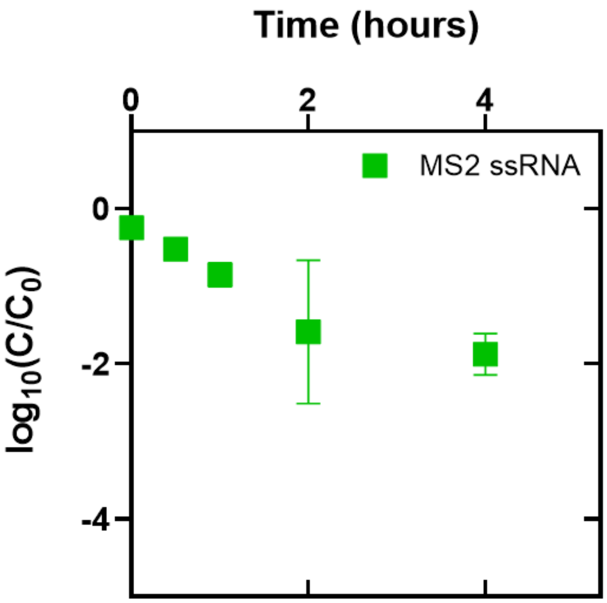


***Figure S3.*** *MS2 ssRNA persistence in untreated wastewater with the addition of RNASecure at 25℃. C represents the concentration of the viral nucleic acids in gene copies µL$^{-1}$ at time t in hours, and $C_0$ represents the initial concentration of the viral nucleic acids at time = 0 based on the amount of stock that was spiked into the wastewater. Initial concentrations in wastewater were ~$10^9$ gc mL$^{-1}$ for MS2 (+)ssRNA. Error bars represent the standard error for three experimental replicates conducted in wastewater samples collected on different days.*

***qPCR assays.*** All DNA and cDNA targets were enumerated for gene copies in each sample using qPCR. Complete assay information for each virus is included in Table S4. For MS2, BCoV, and T4 targets, 20 µL reactions consisted of: 10 µL Biotium Fast EvaGreen Master Mix, 1 µL forward primer at 10mM, 1 µL reverse primer at 10mM, 6 µL nuclease-free water, and 2 µL template. For the T3 target, 20 µL reactions consisted of: 10 µL Biotium Fast EvaGreen Master Mix, 0.8 µL forward primer at 10mM, 0.8 µL reverse primer at 10mM, 0.5 µL ultrapure BSA at 25 mg $mL^{-1}$, 5.9 µL nuclease-free water, and 2 µL template. A no template control (NTC) was included in each qPCR plate for contamination control. All samples and standards were run in triplicate. For each experiment, two additional controls were included and enumerated using qPCR. An inhibition control consisted of a 1:10 dilution of the highest concentrated sample in the wastewater matrix. Gene copies were compared after accounting for the dilution in samples and results are summarized in Figure S4. A background control consisted of extracted RNA or DNA from untreated wastewater used for experiments before spiking viruses into the sample. Targets were measured in wastewater background extracts and results are summarized in Figure S5.

All qPCR reactions were run on an Applied Biosystems QuantStudio™ 3 Real-Time PCR System (Fisher). Prior to running, qPCR plates were centrifuged at 2000xg for 2 minutes to collect the full reaction volume at the bottom of each well and decrease bubbles which may interfere with the assay. Cycle conditions consisted of the following steps: an initial enzyme activation step at 95℃ for 2 minutes followed by 45 cycles of denaturation at 95℃ for 5 seconds, annealing at specified annealing temperature in Table S4 for 5 seconds, an extension step at 72℃ for 25 seconds, a melt curve stage consisting of 95℃ for 15 seconds, 60℃ for 1 minute, and 95℃ for 15 seconds, followed by an indefinite hold at 4℃. Target gene copies were enumerated based on standard curves made from serial dilutions of known quantities of gene targets extracted from the virus stock. The limit of quantification for the assay and efficiency were calculated for analysis and are summarized for each assay in Table S4.

***Table S4.*** *qPCR assays for virus targets*

| Virus Target | Forward Primer and Reverse Primer | Amplicon Length (b or bp) | Annealing Temperature | LOQ[a] | Mean $R^2$ (min to max) | Mean efficiency[b] (min to max) | Ref. |
|---|---|---|---|---|---|---|---|
| MS2 | F: 5’-TGG CAC TAC CCC TCT CCG TAT TCA C-3’<br>R: 5’-GTA CGG GCG ACC CCA CGA TGA C-3’ | 99 b | 60°C | 32.0 | 0.9991<br>(0.9973 to 0.9999) | 91.45<br>(84.17 to 94.98) | This study |
| | F: 5’-GGT TTG ACC TGT GCG AGC TT-3’<br>R: 5’-GTA CGG GCG ACC CCA CGA TGA C-3’ | 268 b | 60°C | 33.4 | 0.9988<br>(0.9982 to 0.9993) | 85.60<br>(80.09 to 89.25) | This study |
| | F: 5’-GGG TCC TGC TCA ACT TCC TG-3’<br>R: 5’-GTA CGG GCG ACC CCA CGA TGA C-3’ | 370 b | 60°C | 34.8 | 0.9938<br>(0.9895 to 0.9978) | 75.47<br>(70.99 to 79.60) | This study |
| BCoV | F: 5’-CTG GAA GTT GGT GGA GTT-3’<br>R: 5’-ATT ATC GGC CTA ACA TAC ATC-3’ | 89 b | 58°C | 32.6 | 0.9959<br>(0.9919 to 0.9992) | 106.53<br>(99.00 to 115.58) | Decaro et al. 2008[3] |
| T3 | F: 5’-CAG TCC AAC TAT GTA CGG AAC AA-3’<br>R: 5’-AAG CGA GTA CCA GAA GCA GTA AT-3’ | 99 bp | 58°C | 30.1 | 0.9992<br>(0.9984 to 0.9998) | 88.98<br>(87.99 to 89.71) | Kettleson et al. 2009[4] |
| T4 | F: 5’-CCA CAA CTA ACC GAG GAA GTA A-3’<br>R: 5’-TGC GAT ATG CTA TGG GTC TTG-3’ | 107 bp | 58°C | 32.2 | 0.9992<br>(0.9986 to 0.9997) | 90.74<br>(89.46 to 92.65) | Lim et al. 2017[5] |

[a]Values are given as cycle threshold (CT)
[b]Efficiency is given as a percentage and was calculated according to the following equation: $E = (10^{(-1/slope)} - 1) \times 100\%$.
LOQ = Limit of quantification

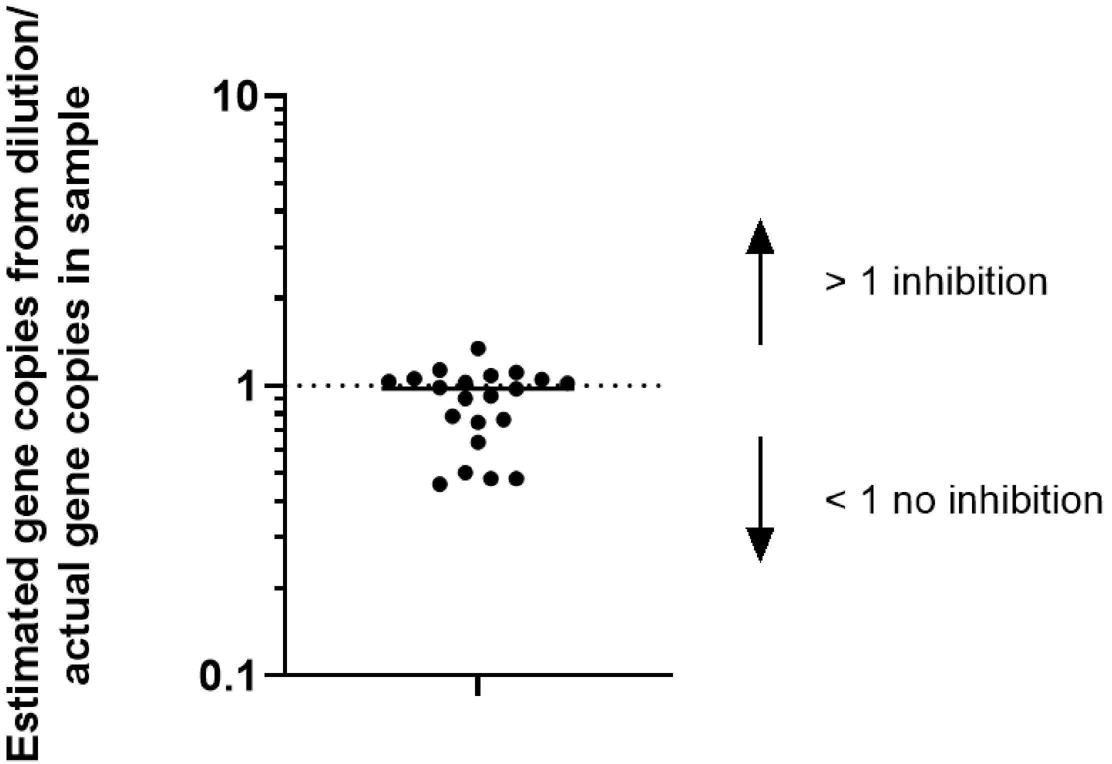


***Figure S4.*** *Comparison of estimated gene copies from diluted sample with actual gene copies in samples. Black line represents the overall mean of the data.*

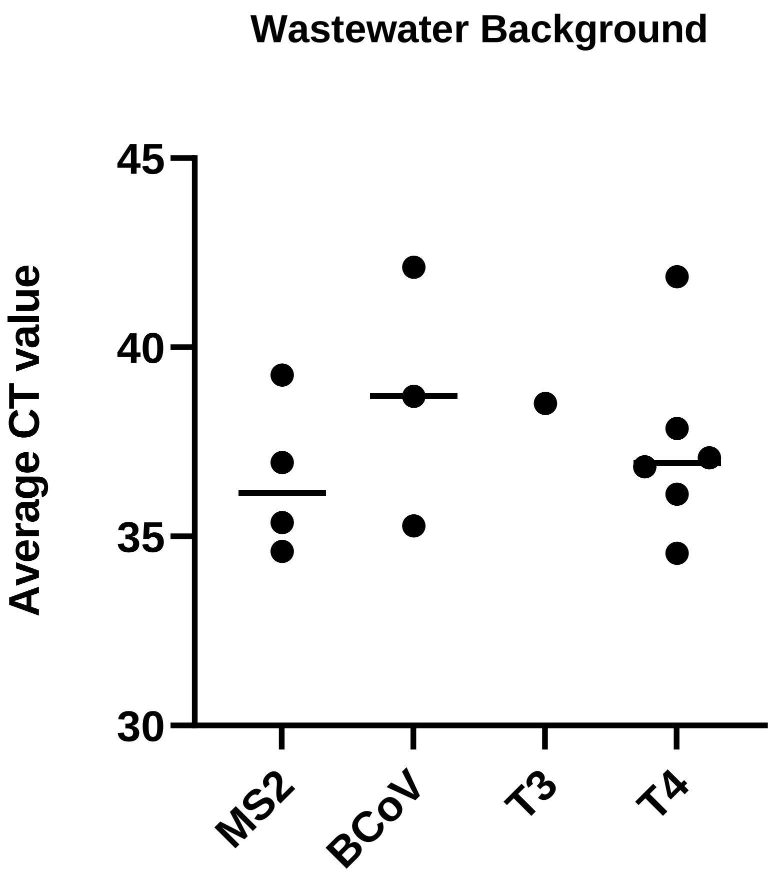


***Figure S5.*** *Measured virus targets in wastewater background. Targets were measured in wastewater extracts without viruses spiked in. Some replicates were not shown since they were below the limit of detection for the assay. Lines represent means of replicates that were above the limit of detection for the assay. In all cases, average CT values were higher in wastewater than in samples and below the limit of quantification.*

***ddPCR assays.*** For each virus stock, ddPCR was used to enumerate absolute quantification of gene copies. Complete assay information for each virus is included in Table S5. For all virus targets, 22 µL reactions consisted of: 11 µL ddPCR Supermix for Probes (No dUTP), 1.1 µL forward primer at 10mM, 1.1 µL reverse primer at 10mM, 1.1 µL probe at 10mM, 4.7 µL nuclease-free water, and 3 µL template. A no template control (NTC) consisting of 3 µL of nuclease-free water was included in each ddPCR plate to detect contamination. Samples were diluted either 1:1000, 1:10000, or 1:100000 based on the assumed number of positive droplets falling within the ideal range of 1000 to 10000. Two dilutions were run for each virus and all diluted samples were run in triplicate.

All ddPCR reactions were loaded onto a Bio-Rad Automated Droplet Generator. Droplet generation was performed with Automated Droplet Generation Oil for Probes (Bio-Rad Cat. No. 1864110). Droplets were formed and deposited onto a plate fitted with an ice block kept at -20℃. All plates were sealed with pierceable foil and loaded onto a Bio-Rad C1000 Touch™ Thermal Cycler. Cycle conditions consisted of the following steps: an initial denaturation at 95℃ for 10 minutes followed by 40 cycles of denaturation at 95℃ for 30 seconds, annealing at 56℃ for 1 minute, and extension at 72℃ for 1 minute, a hold step at 4℃ for 5 minutes, an enzyme deactivation step at 95℃ for 5 minutes, and finally an indefinite hold at 4℃. Finally, droplets were read within 1 hour on a Bio-Rad QX200™ Droplet Reader equipped with Bio-Rad ddPCR Droplet Reader Oil (Bio-Rad Cat. No. 1863004). Target gene copies were enumerated based on the Poisson distribution of positive droplets in the sample. Assay information is summarized in Table S5.

***Table S5.*** *ddPCR assays for virus targets*

| Virus Target | Primers/Probes | Amplicon Length (bp) | Annealing Temperature | Ref. |
|---|---|---|---|---|
| MS2 | Forward: 5’-TGG CAC TAC CCC TCT CCG TAT TCA CG-3’<br>Reverse: 5’-GTA CGG GCG ACC CCA CGA TGA C-3’<br>Probe: 5’-/56-FAM/CAC ATC GAT/ZEN/AGA TCA AGG TGC CTA CAA GC/BHQ_1/-3’ | 99 b | 56°C | Rolfe et al. 2007[6] |
| BCoV | Forward: 5’-CTG GAA GTT GGT GGA GTT-3’<br>Reverse: 5’-ATT ATC GGC CTA ACA TAC ATC-3’<br>Probe: 5’-CCT TCA TAT CTA TAC ACA TCA AGT TGT T-3’ (5’ FAM/ZEN/3’ IBFQ) | 89 b | 56°C | Decaro et al. 2008[3] |
| T3 | Forward: 5’-CCA ACG AGG GTA AAG TGA TAG-3’<br>Reverse: 5’-CGA CGA TAG CGA ATA GGA TAA G-3’<br>Probe: 5’-/HEX/-CC AAC AAC ATC TCT CGC GCA TT-/BHQ_2/-3’ | 351 bp | 56°C | Langenfeld et al. 2021[2] |
| T4 | Forward: 5’-CCA CAA CTA ACC GAG GAA GTA A-3’<br>Reverse: 5’-TGC GAT ATG CTA TGG GTC TTG-3’<br>Probe: 5’-/FAM/-TGC TCC ATC AGA GGA AGA ATG CGA-/BHQ_1/-3’ | 107 bp | 56°C | Lim et al. 2017[5] |

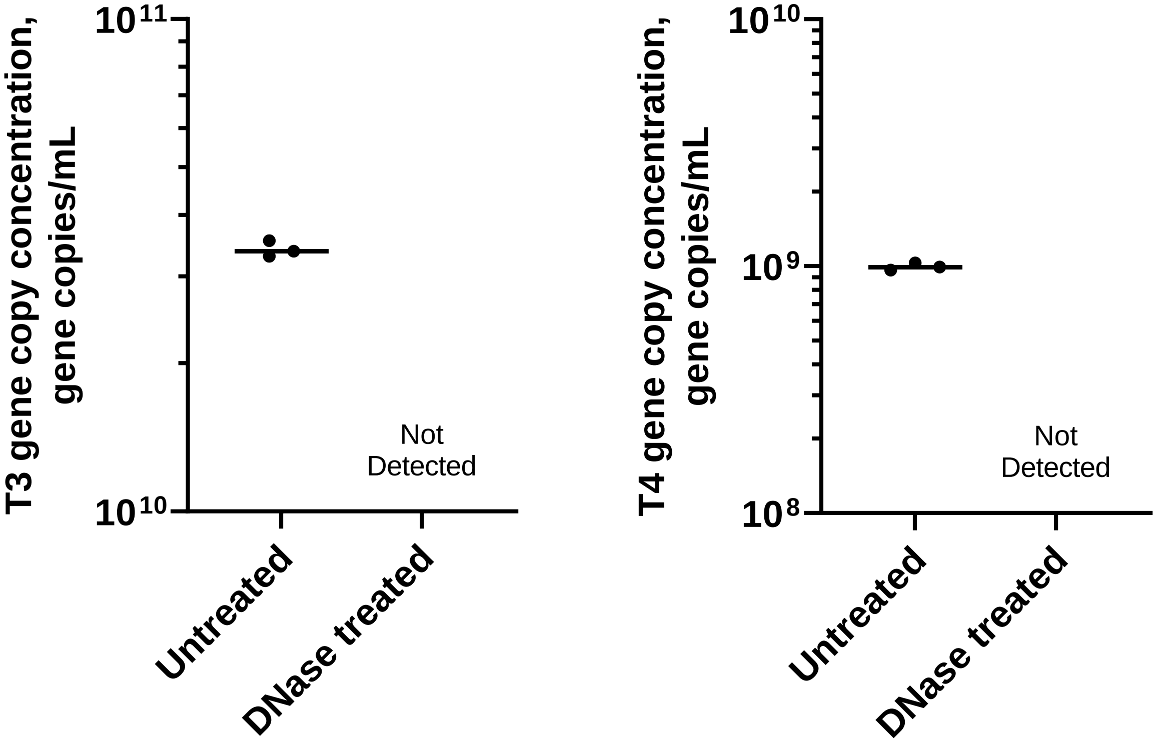


***Figure S6.*** *DNase treatment of T3 and T4 genomes to ensure extraviral nucleic acids. Lines depict the mean of three independent replicates of the same extraviral stock (n = 3). For T3 and T4, extracted dsDNA genomes treated with DNase were not detected in subsequent qPCR reactions and therefore encapsidated genomes were not present after nucleic acid extraction.*

***Table S6.*** *Summary of reported decay rates for encapsidated viral nucleic acids in wastewater*

| Virus target | Genome type and structure | First order decay rate (days$^{-1}$) | $T_{90}$ (days) | Amplicon size | Temperature | Ref. |
|---|---|---|---|---|---|---|
| SARS-CoV-2 | (+)ssRNA, enveloped | 0.183 | 12.6 | 72 b | 25°C | Ahmed et al. 2020[7] |
| SARS-CoV-2 | (+)ssRNA, enveloped | 0.171* | 13.5* | 72 b | 25°C | Ahmed et al. 2020[7] |
| SARS-CoV-2 | (+)ssRNA, enveloped | 0.67 (high titer)$^{+}$ | 3.3$^{+}$ | 113 b | 20°C | Bivins et al. 2020[8] |
| SARS-CoV-2 | (+)ssRNA, enveloped | 0.09 (low titer)$^{+}$ | 26$^{+}$ | 113 b | 20°C | Bivins et al. 2020[8] |
| SARS-CoV-2 | (+)ssRNA, enveloped | 0.84 | 2.74 | 72 b | 20°C | McCall et al. 2021[9] |
| MHV | (+)ssRNA, enveloped | 0.135 | 17.3 | 80 b | 25°C | Ahmed et al. 2020[7] |
| MHV | (+)ssRNA, enveloped | 0.132* | 17.6* | 80 b | 25°C | Ahmed et al. 2020[7] |
| MHV | (+)ssRNA, enveloped | 0.371 | 6.205 | 108 b | 25°C | Chandra et al. 2021[10] |
| Zika Virus | (+)ssRNA, enveloped | 0.11 | 21 | Proprietary (NS1 gene target) | 25°C | Muirhead et al. 2020[11] |
| Zika Virus | (+)ssRNA, enveloped | 0.583 | 3.948 | 66 b | 25°C | Chandra et al. 2021[10] |
| Yellow Fever Virus | (+)ssRNA, enveloped | 0.523 | 4.403 | 89 b | 25°C | Chandra et al. 2021[10] |
| Dengue Virus 2 | (+)ssRNA, enveloped | 0.500 | 4.604 | 78 b | 25°C | Chandra et al. 2021[10] |
| Dengue Virus 3 | (+)ssRNA, enveloped | 0.551 | 4.179 | 74 b | 25°C | Chandra et al. 2021[10] |
| Poliovirus type 2 | (+)ssRNA, naked | NA | NA | 100 b | 19-24°C | Kline et al. 2022[12] |
| Poliovirus type 3 | (+)ssRNA, naked | NA | 7 | 120 b | 19-24°C | Kline et al. 2022[12] |
| Bacteriophage MS2 | (+)ssRNA, naked | 0.17 (first phase) | 5.5 | 99 b | 25°C | This study |
| Bacteriophage T4 | dsDNA, naked | 0.34 (first phase) | 3.3 | 107 bp | 25°C | This study |

*Denotes pasteurized or autoclaved wastewater. $^{+}$Denotes wastewater frozen prior to experiments.

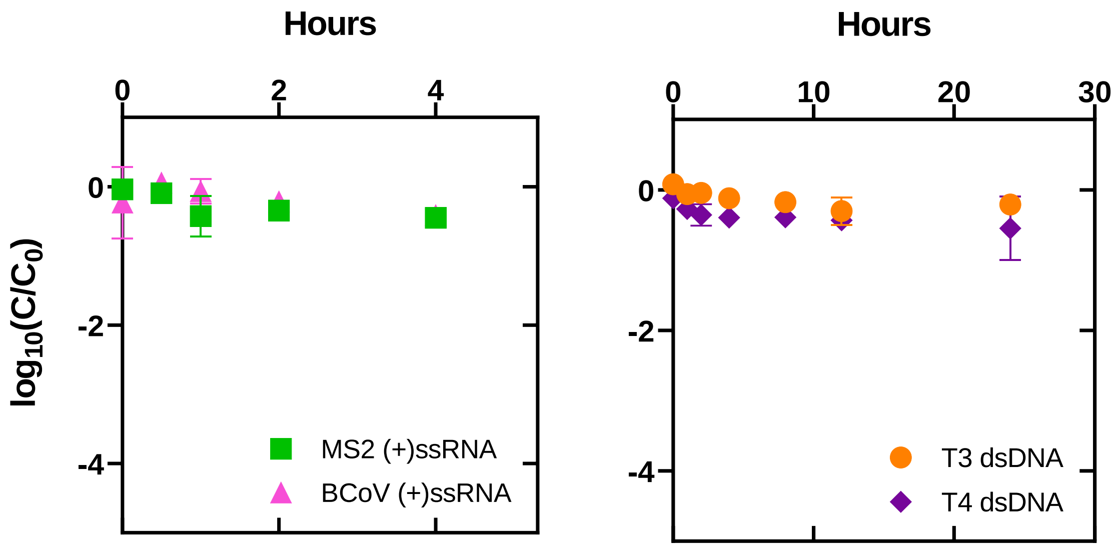


***Figure S7.*** *Extraviral (+)ssRNA and dsDNA decay in PBS at 25℃. C represents the concentration of the viral nucleic acids in gene copies* $\mu L^{-1}$ *at time t in hours and* $C_0$ *represents the initial concentration of the viral nucleic acids at time = 0 based on the amount of stock that was spiked into the wastewater. Initial concentrations in wastewater were* ~$10^9$ *gc* $mL^{-1}$ *for MS2 (+)ssRNA and* ~$10^8$ *gc* $mL^{-1}$ *for BCoV (+)ssRNA, T3 dsDNA, and T4 dsDNA. Open symbols represent measurements that were below the limit of quantification and are plotted as the limit of quantification for each virus assay. Error bars represent the standard error for three experimental replicates conducted in wastewater samples collected on different days.*

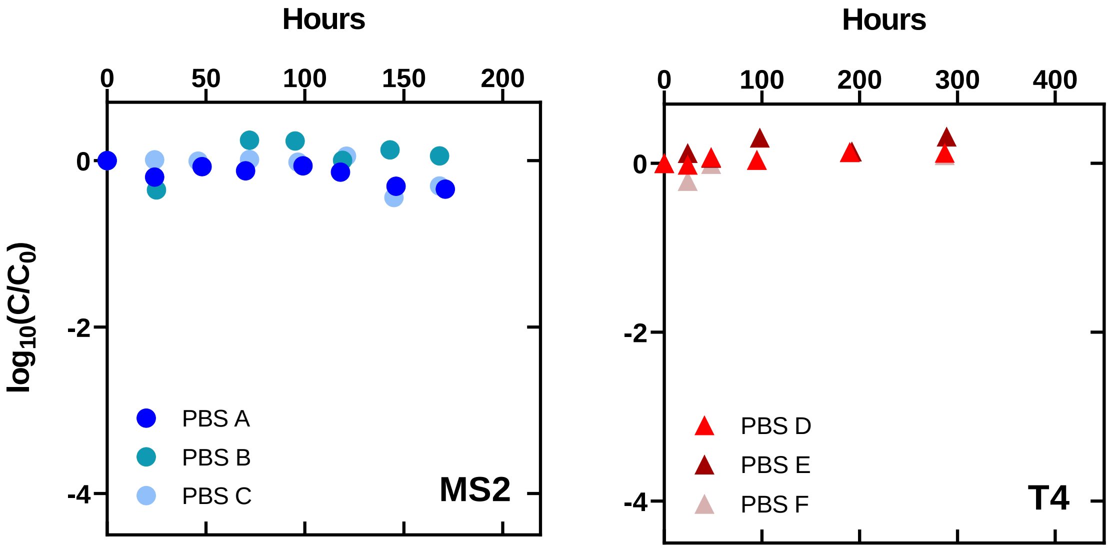


***Figure S8.*** *Encapsidated MS2 (+)ssRNA and T4 dsDNA genome decay in PBS at 25℃. C refers to the concentration of the viral nucleic acids in gene copies μL*$^{-1}$ *at time t in hours.* $C_0$ *represents the initial concentration of the viral nucleic acids at time = 0 based on the amount of gene copies measured immediately after the stocks were added to the sample. Initial concentrations in wastewater were ~*$10^9$ *gc mL*$^{-1}$ *for MS2 and ~*$10^8$ *gc mL*$^{-1}$ *for T4. Open symbols represent measurements that were below the limit of quantification and are plotted as the limit of quantification for each virus assay. Individual experimental replicates are shown for MS2 (PBS A, B, C) and T4 (PBS D, E, F).*

**References**


1. Rockey, N. *et al.* UV Disinfection of Human Norovirus: Evaluating Infectivity Using a Genome-Wide PCR-Based Approach. *Environ. Sci. Technol.* **54**, 2851–2858 (2020).
2. Langenfeld, K., Chin, K., Roy, A., Wigginton, K. & Duhaime, M. B. Comparison of ultrafiltration and iron chloride flocculation in the preparation of aquatic viromes from contrasting sample types. *PeerJ* **9**, e11111 (2021).
3. Decaro, N. *et al.* Detection of bovine coronavirus using a TaqMan-based real-time RT-PCR assay. *J. Virol. Methods* **151**, 167–171 (2008).
4. Kettleson, E. M. *et al.* Airborne Virus Capture and Inactivation by an Electrostatic Particle Collector. *Environ. Sci. Technol.* **43**, 5940–5946 (2009).
5. Lim, S. W., Lance, S. T., Stedman, K. M. & Abate, A. R. PCR-activated cell sorting as a general, cultivation-free method for high-throughput identification and enrichment of virus hosts. *J. Virol. Methods* **242**, 14–21 (2017).
6. Rolfe, K. J. *et al.* An internally controlled, one-step, real-time RT-PCR assay for norovirus detection and genogrouping. *J. Clin. Virol.* **39**, 318–321 (2007).
7. Ahmed, W. *et al.* Decay of SARS-CoV-2 and surrogate murine hepatitis virus RNA in untreated wastewater to inform application in wastewater-based epidemiology. *Environ. Res.* **191**, 110092 (2020).
8. Bivins, A. *et al.* Persistence of SARS-CoV-2 in Water and Wastewater. *Environ. Sci. Technol. Lett.* **7**, 937–942 (2020).
9. McCall, C. *et al.* Modeling SARS-CoV-2 RNA degradation in small and large sewersheds. *Environ. Sci. Water Res. Technol.* **8**, 290–300 (2022).
10. Chandra, F. *et al.* Persistence of Dengue (Serotypes 2 and 3), Zika, Yellow Fever, and Murine Hepatitis Virus RNA in Untreated Wastewater. *Environ. Sci. Technol. Lett.* **8**, 785–791 (2021).
11. Muirhead, A. *et al.* Zika Virus RNA Persistence in Sewage. *Environ. Sci. Technol. Lett.* **7**, 659–664 (2020).
12. Kline, A. *et al.* Persistence of poliovirus types 2 and 3 in waste-impacted water and sediment. *PLOS ONE* **17**, e0262761 (2022).
